# Spike mutations of the SARS-CoV-2 Omicron: more likely to occur in the epitopes

**DOI:** 10.1101/2022.03.03.22271159

**Authors:** Hu Li, Zhiwei Chen, Xiaoqing Liu, Peng Hu

## Abstract

Almost two years since the severe acute respiratory syndrome coronavirus 2 (SARS-CoV-2) outbreak in 2019, and it is still pandemic over the world. SARS-COV-2 continuing to mutate and evolve, which further exacerbated the spread of the epidemic. Omicron variant, as an emerging mutation recently in South Africa, spreaded fastly to other countries worldwide. However, the gene charicterstic of Omicron and the effect on epitopes are still unclear. In this study, we retrieved 800 SARS-CoV-2 full-length sequences from GISAID database on 14 December 2021 (Alpha 110, Beta 101, Gamma 108, Delta 110, Omicron 107, Lambda 98, Mu 101, GH/490R 65). Overall, 1320 amino acid (AA) sites were mutated in these 800 SARS-CoV-2 sequences. Covariant network analysis showed that the covariant network of Omicron variant was significantly different from other variants. Further, 218 of the 1320 AA sites were occurred in the S gene, including 78 high-frequency mutations (>90%). Notably, we identified 25 unique AA mutations in Omicron, which may affect the transmission and pathogenicity of SARS-CoV-2. Finally, we analyzed the effect of Omicron on epitope peptide. As expected, 64.1% mutations (25/39) of Omicron variants were in epitopes, which was significantly higher than in other variants. These mutations may cause a poor response to vaccines to Omicron variants. In conclusion, Omicron variants, as an emerging mutation, should be alerted for us due that it may lead to poor vaccine response, and more data is needed to evaluate the virulence and vaccines responses to this variants.

---

Coronavirus disease 2019 (COVID-19), caused by the severe acute respiratory syndrome coronavirus 2 (SARS-CoV-2) infection, has become a significant global public health threat. So far, over 273 million cases and over 5.3 million deaths have been reported globally. The SARS-CoV-2 variant B.1.1.529 in South Africa was first reported to the World Health Organization (WHO) on 24 November 2021. It appears to be rapidly spreading, and the WHO classified it as a variant of concern (VOC) designating it as Omicron. The Omicron variant has been reported in travel-related cases in several European countries and Australia, Brazil, Canada, Hong Kong, Israel, Japan, Nigeria, Norway, Sweden, and the United Kingdom.

In this study, 6,092,003 sequences of the SARS-CoV-2 were retrieved from GISAID^1^ on 14 December 2021 (https://www.gisaid.org/). To the more convenient analysis of the mutation patterns among different variants, 800 latest high-coverage complete genomes SARS-CoV-2 strains (Alpha 110, Beta 101, Gamma 108, Delta 110, Omicron 107, Lambda 98, Mu 101, GH/490R 65) were included in this study and listed in Supplementary Table1. The complete genome sequences were aligned with the reference genome of SARS-CoV-2 (NC_045512) by MAFFT (version 7.0, https://mafft.cbrc.jp/alignment/server/). The aligned genomes were then edited manually according to the reference sequence by BioEdit software. Finally, a total of 1320 amino acid (AA) sites in all open reading frames (ORFs) were detected mutations from all the 800 sequences (Figure 1A). Of these mutations, 791 (59.92%) was in ORF1ab, followed by ORF2 (218, 16.52%), ORF9 (99, 7.50%), ORF3 (88, 6.67%), the mutations in other ORFs were infrequent (all < 5%).

**Figure 1.**
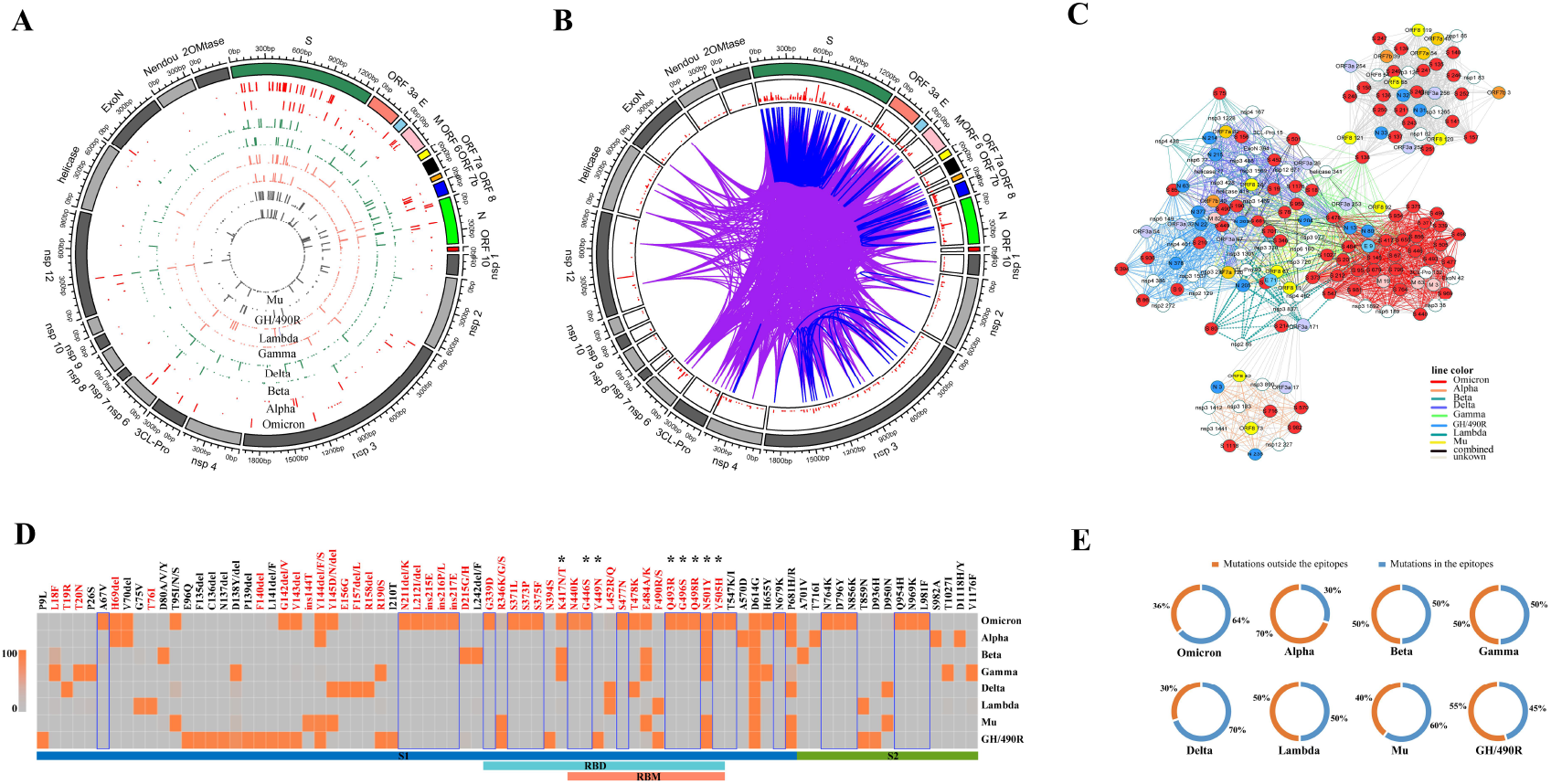
The characteristic of the SARS-CoV-2 mutations. (A) Genome-wide amino acid mutation rates of different variants of SARS-CoV-2. (B) Plotting of predicted covariant and interacting residues in clinical strains. 2268 covariant residue pairs were detected in the 161 isolates. Covarying pairs in the same region were shown in blue, and pairs in the different regions were shown in purple. (C) Amino acid covariant network graph. The node colour indicates the viral genomic segment. (D) Common AA mutations in spike protein distributed among Omicron and other variants. blue box: unique AA mutations in the Omicron variant; *: key AA mutations in the RBD region interface with ACE2; red font: AA mutations in the 163 epitopes. (E) Percentage of mutations in the 163 epitopes in Omicron and other variants. AA: amino acid, RBD: receptor binding domain, RBM: receptor binding motif, ACE2: Angiotensin-Converting Enzyme 2.

The protein-coding region of SARS-CoV-2 was used to analyze covariant pairs of amino acid position. Covarying positions were identified using OMES algorithms^2^. And an OMES score of 3 was used as the cutoff for analyses. Amino-acid covariant networks were visualized by Cytoscape version 3.7.2. Sequences enrolled in this study were used to analyze covarying pairs according to different variants (Figure S1). Significant differences in covariant residue networks among different SARS-CoV-2 variants were observed. Then we identified 2268 covarying amino acid pairs most highly represented in 161 isolates (including 20 randomly selected isolates of each subtype and a reference sequence) and found these form small closed networks within and between protein. Amino acid covarying network in the SARS-CoV-2 shown in Figure 1B, which revealed that several mutations in the spike protein are predicted to be covariant in our 161 strains with residues in the ORF1ab, E, M, N, ORF3a, ORF6, ORF7, ORF8, which suggest there is an evolutionary relationship between these areas. And from Figure 1C, We observed Omicron variant has significant differences in covariant residue networks compared with other SARS-COV-2 variants.

Considering the critical function of spike protein in virus entry and antibody response, we focus on the diversity of spike protein among the different variants. Among the 218 AA mutations in the spike, 78 common mutations (high frequency [> 90%] occurred in each variant) were visualized by heatmap. As shown in Figure 1D, the diversity of mutations in each variant was different. D614G, enhanced virus stability, infectivity and transmission^3,4^, was observed in all variants’ strains. Notably, 25 unique AA mutation sites were observed in the Omicron, including one deletion mutation (L212 del) and three insertion mutations (ins215-217 EPE). Of the 25 AA mutations, 11 were presented in the receptor binding domain (RBD) region (G339D, S371L, S373P, S375F, N440K, G446S, S477N, Q493R, G496S, Q498R and Y505H), and 7 occurred in the receptor-binding motif (RBM) region. For the 17 residues in RBD interfaces with Angiotensin-Converting Enzyme 2 (ACE2)^5^, eight were mutated in the different variants. In detail, K417N was observed in the Omicron, Beta and Gamma variants, which reduces ACE2 binding significantly^6,7^. Whereas N501Y, which exhibit increased ACE2 binding affinity^8,9^, was detected in all other 6 variants except Delta and Lambda. In addition, five residues only occurred in the Omicron, G446S, Q493R, G496S, Q498R and Y505H. The Cryo-EM structure of spike protein of Omicron variant showed that new salt bridges and hydrogen bonds formed by mutated residues R493, S496 and R498 in the RBD with ACE2 likely make spike-ACE2 binding stronger, which appear to compensate other Omicron mutations such as K417N known to reduce ACE2 binding affinity^10^. However, a recent in vitro study11 showed that the Omicron variant exhibited reduced infectivity in human lung epithelia-derived CaLu-3 cells compared to other variants.

To further describe the relationship between the spike mutations and epitopes of the SARS-CoV-2, a total of 163 reported epitopes were downloaded from the Immune Epitope Database (IEDB) (http://www.iedb.org/) (Supplementary Table2). Among these epitopes, 161 are discontinuous, and 2 are linear epitopes. 52.6% (41/78) of spike mutations are located within the epitopes (Figure 1D). Especially in the Omicron, 64.1% (25/39) of the mutations are located within the epitopes, which was significantly higher than other variants except for the Delta (Figure 1E). 64.0% (16/25) of unique AA mutations in Omicron were in the epitopes, whereas the previous VOCs displayed mutations were only partially (47.2%, 25/53) located within the epitope. All (107/107) of the Omicron strains have at least one epitope mutation. The mutations in the Omicron strains might impact recognition of this domain by neutralizing antibodies of vaccine-induced immunity resulting in immune escape.

Hene, more laboratory and epidemiological studies are warranted to assess the impact of the Omicron variant on vaccine effectiveness and breakthrough infections, including in individuals who have received booster doses.

## Data Availability

https://www.gisaid.org/

## Supplementary

**Supplementary Figure 1**. Genome-wide amino acid covariant network of different variants of the SARS-CoV-2.

**Supplementary Table 1**. List of the SARS-CoV-2 strains retrieved from the GISAID database used in this study.

**Supplementary Table 2**. The reported epitopes of the SARS-CoV-2 spike.

