## Supplementary figures and images for "Spike mutations of the SARS-CoV-2 Omicron: more likely to occur in the epitopes"

### Supplementary Figure 1

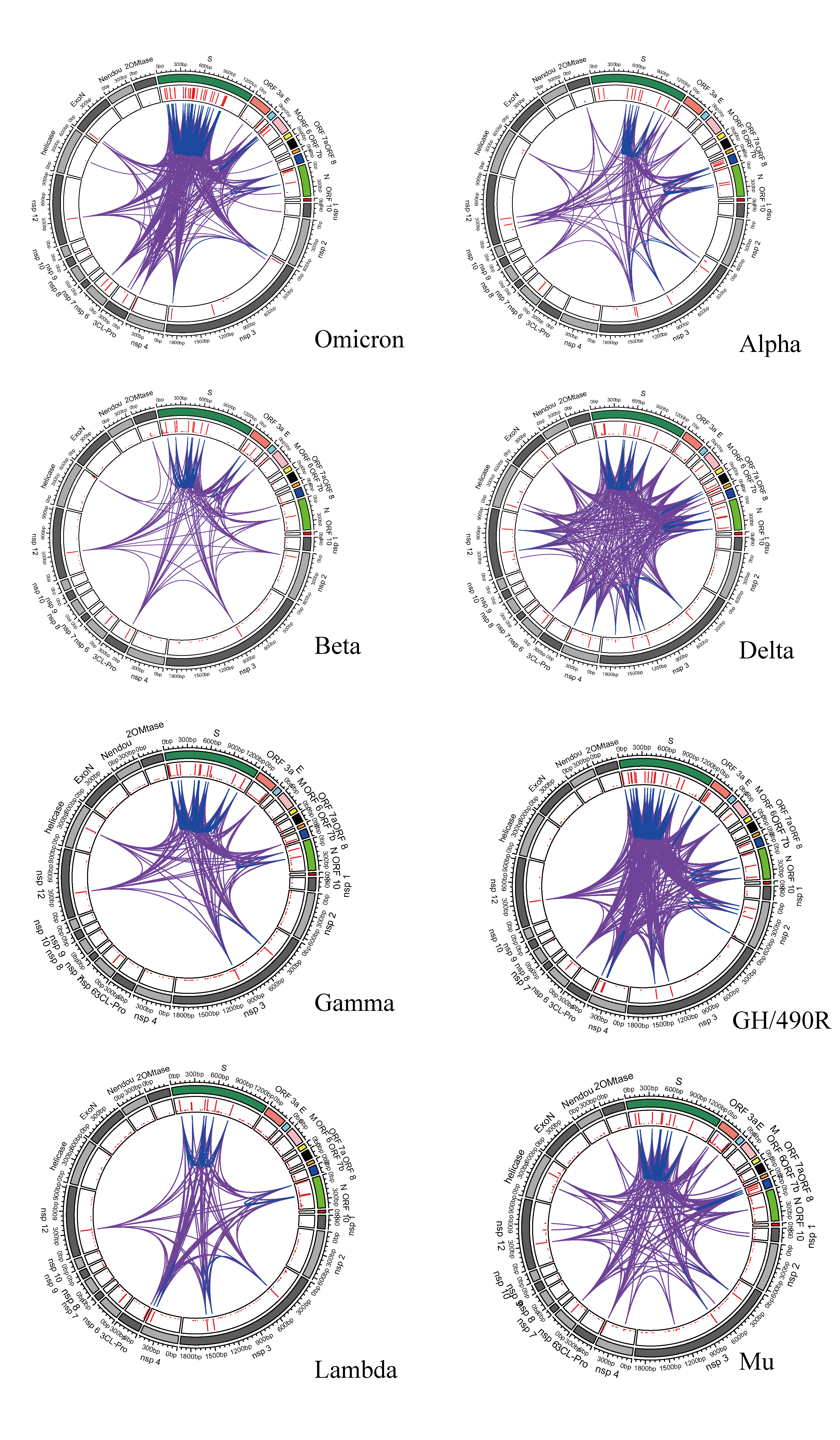
