## Supplementary Table 1 for "Spike mutations of the SARS-CoV-2 Omicron: more likely to occur in the epitopes"

We gratefully acknowledge the following Authors from the Originating laboratories responsible for obtaining the specimens, as well as the Submitting laboratories where the genome data were generated and shared via GISAID, on which this research is based.

All Submitters of data may be contacted directly via www.gisaid.org

Authors are sorted alphabetically.

| Accession ID | Originating Laboratory | Submitting Laboratory | Authors |
|---|---|---|---|
| EPI_ISL_5887322 | ADPH | ADPH | Adrianna Walker |
| EPI_ISL_5914875 | AREA DE SALUD BARVA (COOPESIBA) | Inciensa, Instituto Costarricense de Investigación y Enseñanza en Nutrición y Salud | Adriana Godínez; Claudio Soto-Garita; Estela Cordero; Francisco Duarte; Hebleen Porras; José Luis Vargas; Mariela Gutiérrez; Melany Calderón; Sofia Herrera & Angélica Espinoza Fontana |
| EPI_ISL_6492078 | AULSS 8 Berica | Istituto Zooprofilattico Sperimentale delle Venezie | Adelaide Milani; Alessia Schivo; Alice Fusaro; Ambra Pastori; Angela Salomoni; Annalisa Salviato; Antonia Ricci; Calogero Terregino; Edoardo Giussani; Elisa Palumbo; Erika Giorgia Quaranta; Isabella Monne |
| EPI_ISL_4080668 | AUSTRAL-omics, UACh | AUSTRAL-omics, UACh | Andrea Silva; Carolina Encina; Cristian Molina; Daniela Plaza; Luis Guzmán; Suany Quesada |
| EPI_ISL_5860239, EPI_ISL_6513874, EPI_ISL_7412265 | Aegis Sciences Corporation | Centers for Disease Control and Prevention Division of Viral Diseases, Pathogen Discovery | Alec Vest; Benjamin Rambo-Martin; Christopher Gulvick; Clinton Paden; Cyndi Clark; Dakota Howard; Dhwani Batra; Dillon Nall; Duncan MacCannell; Erisa Sula; Ethan Sanders; Holly Houdeshell; Jason Caravas; Kristine Lacek; Matthew Hardison; Matthew Schmerer; Ola Kvalvaag; Patrick Campbell; Peter Cook; Rob Case; Scott Sammons; Shatavia Morrison; Shaun Westlund; Tymeckia Kendall; Victoria Caban Figueroa; Vikramsinha Ghorpade; Yvette Unoarumhi |
| EPI_ISL_6914018, EPI_ISL_6914029, EPI_ISL_6914031 | Ampath Laboratories | National Institute for Communicable Diseases of the National Health Laboratory Service | Amoako DG; Bhiman JN; Everatt J; Ismail A; Mahlangu B; Mnguni A; Mohale T; Ntuli N; Scheepers C; Wolter N |
| EPI_ISL_7338921 | Azienda Sanitaria dell´Alto Adige - Laboratorio Aziendale di Microbiologia e Virologia | Azienda Sanitaria dell'Alto Adige | Irene Bianconi |
| EPI_ISL_6248670 | BIO-REFERENCE LABORATORIES | Wadsworth Center, New York State Department of Health | Alexis Russell; Catharine Prussing; Daryl M. Lamson; Erasmus Schneider; Erica Lasek-Nesselquist; John Kelly; Jonathan Plitnick; Kirsten St. George; Matthew Shudt; Melissa A Leisner; Navjot Singh |
| EPI_ISL_7570539, EPI_ISL_7570553, EPI_ISL_7570555 | Bluewater Diagnostic Laboratory, LLC | University of Louisville Sequencing Technology Center | Alonzo Shepherd; Aryan Neupane; Elizabeth Hudson; Eric C. Rouchka; Erik Korte; Julia H. Chariker; Melissa L. Smith; William Lauer |
| EPI_ISL_7380512, EPI_ISL_7380515 | Botswana Harvard HIV Reference Laboratory | Botswana Harvard HIV Reference Laboratory | Boitumelo Zuze; Botshelo Radibe; Dorcas Maruapula; Joseph Makhema; Keoratile Ntshambiwa; Kgomotso Moruisi; Legodile Kooepile; Mosepele Mosepele; Mphaphi B. Mbulawa; Ontlametse T. Bareng; Pamela Smith-Lawrence; Roger Shapiro; Sefetogi Ramaologa; Shahin Lockman; Sikhulile Moyo; Simani Gaseitsiwe; Thongbotho Mphoyakgosi; Wonderful T. Choga |
| EPI_ISL_6751417, EPI_ISL_6751445, EPI_ISL_7314617 | British Columbia Centre For Disease Control | BCCDC Public Health Laboratory | Ana Pacagnella; Corrinne Ng; Dan Fornika; John Tyson; Kim Macdonald; Kimia Kamelian; Linda Hoang; Loretta Janz; Mel Krajden; Prystajecky Natalie; Robert Azana; Shannon Russell |
| EPI_ISL_2825823, EPI_ISL_2825959, EPI_ISL_2921738 | CDPH VBL | California Department of Public Health | CDPH-COVIDNet; UCLA Technology Center for Genomics & Bioinformatics |
| EPI_ISL_7427856 | CENTOGENE Frankfurt Laboratory: Niederlassung Industriepark Höchst | Robert Koch Institute | |
| EPI_ISL_6967056, EPI_ISL_6967089, EPI_ISL_6967130, EPI_ISL_6967137 | CH SAINT-PHILIBERT | CHU Lille - Laboratoire de Virologie | AIT YAHYA Emilie; ALIDJINOU Enagnon Kazali; BOCKET Laurence; DEMAY Christophe; ENGELMANN Ilka; GUIGON Aurélie; LAMBERT Valérie; LAZREK Mouna; LOMBART Marion; NOBILLIAUX Florian; VEYER Nathalie |
| EPI_ISL_6779162, EPI_ISL_6779495, EPI_ISL_6779496 | CHU - Saint-Denis | Laboratoire de virologie, CNR arbovirus Associé, Chu de la Réunion | Anne-Julie Gourdé; Etienne Frumence; Marie-Christine Jaffar Bandjee; Nicolas Traversier; Rubens Lhonneur; Sabrina Petit Genet |
| EPI_ISL_6779500 | CHU - Saint-Pierre | Laboratoire de virologie, CNR arbovirus Associé, Chu de la Réunion | Anne-Julie Gourdé; Etienne Frumence; Marie-Christine Jaffar Bandjee; Nicolas Traversier; Rubens Lhonneur; Sabrina Petit Genet |
| EPI_ISL_6967061, EPI_ISL_6967069, EPI_ISL_6967080, EPI_ISL_6967097, EPI_ISL_6967104, EPI_ISL_6967111, EPI_ISL_6967116, EPI_ISL_6967125 | | | |
| see above | CHU LILLE | CHU Lille - Laboratoire de Virologie | AIT YAHYA Emilie; ALIDJINOU Enagnon Kazali; BOCKET Laurence; DEMAY Christophe; ENGELMANN Ilka; GUIGON Aurélie; LAMBERT Valérie; LAZREK Mouna; LOMBART Marion; NOBILLIAUX Florian; VEYER Nathalie |
| EPI_ISL_7609283 | CHU TOULOUSE | CNR Virus des Infections Respiratoires - France SUD | Antonin Bal; Bruno Lina; Bruno Simon; Gregory Destras; Gwendolyne Burfin; Hadrien Regue; Laurence Josset; Martine Valette; Quentin Semanas |
| EPI_ISL_7507406, EPI_ISL_7507515, EPI_ISL_7507685, EPI_ISL_7508821 | Cedars-Sinai Medical Center, Molecular Pathology Laboratory of Department of Pathology & Laboratory Medicine and Genomic Core | Cedars-Sinai Medical Center, Molecular Pathology Laboratory of Department of Pathology & Laboratory Medicine and Genomic Core | Brian Davis; Eric Vail; Jasmine T Plummer; Jorge Mario Sincuir Martinez; Stephanie Chen; Wenjuan Zhang |
| EPI_ISL_7019047 | Charité Universitätsmedizin Berlin, Institute of Virology | Charité Universitätsmedizin Berlin, Institute of Virology | Barbara Mühlemann; Christian Drosten; Julia Schneider; Julia Tesch; Jörn Beheim-Schwarzbach; Talitha Veith; Terry Jones; Tobias Bleicker; Victor M Corman |
| EPI_ISL_7337468 | Charlotte Maxeke Johannesburg Academic Hospital | National Institute for Communicable Diseases of the National Health Laboratory Service | Amoako DG; Bhiman JN; Everatt J; Ismail A; Mahlangu B; Mnguni A; Mohale T; Ntuli N; Scheepers C |
| EPI_ISL_7337527, EPI_ISL_7337528 | Chris Hani Baragwanath Laboratory | National Institute for Communicable Diseases of the National Health Laboratory Service | Amoako DG; Bhiman JN; Everatt J; Ismail A; Mahlangu B; Mnguni A; Mohale T; Ntuli N; Scheepers C |
| EPI_ISL_7266056 | Clinical Microbiology Laboratory, Tel Aviv Sourasky Medical Center | Clinical Microbiology Laboratory, Tel Aviv Sourasky Medical Center | Alon Ziv; Amos Adler; Katya Levytskyi; Lior Handler; Ora Halutz |
| EPI_ISL_7462438 | Cliniques universitaires Saint-Luc | UCLouvain/IREC/MBLG-CTMA | Benoit Kabamba Mukadi; Bertrand Bearzatto; Jean-Luc Gala; Nicolas Pinte; Paul Blanpain; Simon Ophélie; Valentin Coste |
| EPI_ISL_6951145 | Color Genomics | Chiu Laboratory, University of California, San Francisco | Alicia Sotomayor-Gonzalez; Alicia Zhou; Amy Garlin; Charles Chiu; Darpun Sachdev; Katherine Hernandez; Scott Topper; Susan Philip; Venice Servellita; Yueyuan Zhang |
| EPI_ISL_7451089 | Cruz Vermelha Portuguesa | Instituto Nacional de Saude (INSA) | Borges et al |
| EPI_ISL_4405189 | Departamento de Biología y genética molecular, IACA Laboratorios. | Área de Secuenciación del Laboratorio de Virología del Hospital de Niños Dr. Ricardo Gutierrez on behalf of 'Proyecto Argentino Interinstitucional de genomica de SARS-CoV-2' (PAIS Consortium) | ; A; Acuña; D; Goya; LE; Lusso; MI; MS; Masciovecchio MV; Nabaes Jodar; Natale; S; Streitenberger ER; Suárez; Valinotto; Viegas, M. |
| EPI_ISL_6841980, EPI_ISL_6841981, EPI_ISL_7138045, EPI_ISL_7357684, EPI_ISL_7385702 | Department of Microbiology, The University of Hong Kong | Department of Microbiology, The University of Hong Kong | Kelvin K.W. To; Kwok-Yung Yuen |
| EPI_ISL_7201444 | Department of Virology and Immunology, | Department of Virology, Faculty of Medicine, University of Helsinki, | Hanna Jarva; Hanna Liimatainen; Hanna Vauhkonen; Hussein Alburkat; Maija Lappalainen; Mert Erdin; Olli Vapalahti; Phuoc Truong; Ravi Kant; Sari Hannula; Satu Kurkela; Teemu Smura |

| | | | |
|---|---|---|---|
| | University of Helsinki and Helsinki University Hospital, Huslab Finland | Helsinki, Finland | |
| EPI_ISL_6959926, EPI_ISL_6959935 | Division of Emerging Infectious Diseases, Bureau of Infectious Diseases Diagnosis Control, Korea Disease Control and Prevention Agency | Division of Emerging Infectious Diseases, Bureau of Infectious Diseases Diagnosis Control, Korea Disease Control and Prevention Agency | Ae Kyung Park; Chae Young Lee; Eun-Jin Kim; Hyuck Jin Lee; Il-Hwan Kim; Jeong-Ah Kim |
| EPI_ISL_2093805 | Dutch COVID-19 response team | National Institute for Public Health and the Environment (RIVM) | Adam Meijer; AnneMarie van den Brandt; Annelies Kroneman; Bas van der Veer; Chantal Reusken; Dennis Schmitz; Dirk Eggink; Euníce Then; Florian Zwagemaker; Harry Vennema; James Groot; Jeroen Cremer; Karim Hajji; Kim Freriks; Linda van de Nes; Lisa Wijsman; Lynn Aarts; Melissa van Tuil; Ryanne Jaarsma; Sanne Bos; Sharon van den Brink; Sjoerd Kuiling; on behalf of the national COVID-19 response team |
| EPI_ISL_7160042 | Dynacare | National Microbiology Laboratory (NML) | Anna Majer; Anneliese Landgraff; CanCOGeN's metadata curation team; Darian Hole; Dynacare Brampton COVID-19 Diagnostic team; Elsie Grudeski; Gary Van Domselaar; Gordon Jolly; Grace Seo; Jennifer Tanner; Madison Chapel; Morag Graham; Natalie Knox; Nathalie Bastien; Philip Mabon; Public Health Agency of Canada CanCOGeN team; Rhiannon Huzarewich; Russell Mandes; Shari Tyson; Timothy Booth; Yan Li |
| EPI_ISL_7590504 | EXALAB LE HAILLAN | CNR Virus des Infections Respiratoires - France SUD | Antonin Bal; Bruno Lina; Bruno Simon; Gregory Destras; Gwendolyne Burfin; Hadrien Regue; Laurence Josset; Martine Valette; Quentin Semanas |
| EPI_ISL_6721823, EPI_ISL_6721832 | Evandro Chagas Institute | Evandro Chagas Institute | Delana Andreza Melo Bezerra; Dielle Monteiro Teixeira; Jedson Ferreira Cardoso; Jessylene de Almeida Ferreira; Kenny da Costa Pinheiro; Luana Soares Barbagelata; Luana da Silva Soares; Mirleide Cordeiro dos Santos; Patrícia dos Santos Lobo; Rayssa Layna da Silva Bedran; Sandro Patroca da Silva |
| EPI_ISL_2860404, EPI_ISL_5925264, EPI_ISL_5925266, EPI_ISL_5925267, EPI_ISL_5925268, EPI_ISL_5925269, EPI_ISL_5925270, EPI_ISL_5925271, EPI_ISL_5925272, EPI_ISL_5925274, EPI_ISL_5925275, EPI_ISL_5925277, EPI_ISL_5925278, EPI_ISL_5925282, EPI_ISL_5925284, EPI_ISL_5925285, EPI_ISL_5925286, EPI_ISL_5925287, EPI_ISL_5925288, EPI_ISL_5925290, EPI_ISL_5925291, EPI_ISL_5925292, EPI_ISL_5925293, EPI_ISL_5925294, EPI_ISL_5925295, EPI_ISL_5925296, EPI_ISL_5925297, EPI_ISL_5925300, EPI_ISL_5925306, EPI_ISL_5925310, EPI_ISL_5925311, EPI_ISL_7313687 | | | |
| see above | Florida Bureau of Public Health Laboratories | Florida Bureau of Public Health Laboratories | Jason Blanton; Namratha Tarigopula; Sarah Schmedes; Tiffany Splatt |
| EPI_ISL_6701387, EPI_ISL_6701390, EPI_ISL_6701391, EPI_ISL_6701394, EPI_ISL_6701396, EPI_ISL_6701397, EPI_ISL_6701398, EPI_ISL_6701400, EPI_ISL_6701401, EPI_ISL_6701403, EPI_ISL_6701405, EPI_ISL_6701406, EPI_ISL_6701407, EPI_ISL_6701408, EPI_ISL_6701410, EPI_ISL_6701411, EPI_ISL_6701412, EPI_ISL_6701418 | | | |
| see above | Fundação Ezequiel Dias | Instituto René Rachou / Fiocruz Minas | André Bernardes; Anna Salim; Caroline Penido; Felipe Iani; Flávio Araújo; Gabriel Fernandes; Pedro Alves; Rubens do Monte Neto; Thaís Silva |
| EPI_ISL_2508582, EPI_ISL_2557281, EPI_ISL_2756227, EPI_ISL_3537150, EPI_ISL_3987632, EPI_ISL_4005199, EPI_ISL_5879952, EPI_ISL_5879997, EPI_ISL_5879998, EPI_ISL_5880137, EPI_ISL_5880147, EPI_ISL_5880163, EPI_ISL_5880205, EPI_ISL_5880206, EPI_ISL_5880215, EPI_ISL_5880275, EPI_ISL_5880312, EPI_ISL_5880319 | | | |
| see above | Genetica Molecular and Subdepartamento de Virologia ISP Chile | Instituto de Salud Publica de Chile | Andres Castillo; Barbara Parra; Constanza Campano; Gisselle Barra; Javier Tognarelli; Jorge Fernandez; Karen Orostica; Loredana Arata; Patricia Bustos; Rodrigo Fasce; Soledad Ulloa |
| EPI_ISL_6892639 | Germano de Sousa | Instituto Nacional de Saude (INSA) | Borges et al |
| EPI_ISL_7569461, EPI_ISL_7569502, EPI_ISL_7569529, EPI_ISL_7569530, EPI_ISL_7569532, EPI_ISL_7569533, EPI_ISL_7569535, EPI_ISL_7569536, EPI_ISL_7569538, EPI_ISL_7569542, EPI_ISL_7569547, EPI_ISL_7569548, EPI_ISL_7569551, EPI_ISL_7569554, EPI_ISL_7569555, EPI_ISL_7569556, EPI_ISL_7569557, EPI_ISL_7569558, EPI_ISL_7569561, EPI_ISL_7569562, EPI_ISL_7569563, EPI_ISL_7569564, EPI_ISL_7569568, EPI_ISL_7569569, EPI_ISL_7569571, EPI_ISL_7569572, EPI_ISL_7569574, EPI_ISL_7569575, EPI_ISL_7569577, EPI_ISL_7569578, EPI_ISL_7569579, EPI_ISL_7569580, EPI_ISL_7569582, EPI_ISL_7569583, EPI_ISL_7569585, EPI_ISL_7569587, EPI_ISL_7569588, EPI_ISL_7569589, EPI_ISL_7569590, EPI_ISL_7569591 | | | |
| see above | Gravity Diagnostics, LLC | University of Louisville Sequencing Technology Center | Aryan Neupane; Elizabeth Hudson; Eric C. Rouchka; James P. Canner; Julia H. Chariker; Melissa L. Smith; Ryan Walker; William Lauer |
| EPI_ISL_6913917 | Grupo CR Diagnosticos | Instituto Adolfo Lutz Strategic Laboratory | Claudio Tavares Sacchi; Karoline Rodrigues Campos |
| EPI_ISL_6226805 | HOSPITAL UNIVERSITARIO 12 DE OCTUBRE | HOSPITAL UNIVERSITARIO 12 DE OCTUBRE | Carmen Martín-Higuera; Esther Viedma; Irene Múñoz-Gallego; M.ª Dolores Folgueira; Mar Aguilera; Noelia Moral; Rafael Delgado; Sagrario Zurita |
| EPI_ISL_5886781 | HOSPITAL UNIVERSITARIO CENTRAL DE ASTURIAS | Laboratorio de Virología HUCA | ; Alba L; Alvarez-Arguelles ME; Boga JA; Costales I; Coto E; González-Alba JM; Gómez de Oña J; Martín-Rodríguez G; Melón S; Perez-Martínez Z; Rojo S; Sandoval M |
| EPI_ISL_7337440 | Helen Joseph Laboratory | National Institute for Communicable Diseases of the National Health Laboratory Service | Amoako DG; Bhiman JN; Everatt J; Ismail A; Mahlangu B; Mnguni A; Mohale T; Ntuli N; Scheepers C |
| EPI_ISL_5947562 | Helix | Centers for Disease Control and Prevention Division of Viral Diseases, Pathogen Discovery | Benjamin Rambo-Martin; Christopher Gulvick; Clinton Paden; Dakota Howard; Dhwani Batra; Duncan MacCannell; Erisa Sula; Helix CA; Jason Caravas; Kristine Lacek; Matthew Schmerer; Peter Cook; Scott Sammons; Shatavia Morrison; Tymeckia Kendall; Victoria Caban Figueroa; Yvette Unoarumhi |
| EPI_ISL_6590782 | Home Quarantine Taskforce | Hong Kong Department of Health | Alan K.L. Tsang; Edman T.K. Lam; Ken H.L. Ng; Peter C.W. Yip; Rickjason C.W. Chan |
| EPI_ISL_7268331, EPI_ISL_7588318, EPI_ISL_7588324 | Hopital | National Reference Center for Viruses of Respiratory Infections, Institut Pasteur, Paris | Angela Brisebarre; Camille Capel; Christophe Malabat; Corinne Maufrais; Cécile FARRUGIA; Etienne Simon-Lorière; Frédéric Lemoine; Julien Fumey; Louise Lefrançois; Marion Barbet; Maud Vanpeene; Méline Bizard; Pierre PATOZ; Slim El Khiari; Stéphanie VAN AGT; Sylvie Behillil; Sylvie Van der Werf; Vincent Enouf |
| EPI_ISL_6471387 | Hospital | National Reference Center for Viruses of Respiratory Infections, Institut Pasteur, Paris | Angela Brisebarre; Anne-France Georgel; Camille Capel; Christophe Malabat; Corinne Maufrais; Etienne Simon-Lorière; Frédéric Lemoine; Julien Fumey; Louise Lefrançois; Marion Barbet; Maud Vanpeene; Méline Bizard; Slim El Khiari; Sylvie Behillil; Sylvie Van der Werf; Vincent Enouf |
| EPI_ISL_6718313, EPI_ISL_6718389 | Hospital Universitari Dr. Josep Trueta | Institut d'Investigació Biomèdica de Girona \| Hospital Universitari Dr. Josep Trueta | Bernat del Olmo; Mel·lina Pinsach; Meritxell Deulofeu; Nuria Esther Neto; Paula Costa |
| EPI_ISL_6268734 | Hospital Universitari Vall d'Hebron - Vall d'Hebron Institut de Recerca | Hospital Universitari Vall d'Hebron - Vall d'Hebron Institut de Recerca | Alejandra González-Sánchez; Andrés Antón; Ariadna Rando; Carla Castillo; Cristina Andrés; Damir Garcia-Cehic; Josep Quer; Juliana Esperalba; Karen García; Maria Carmen Martin; Maria Gema Codina; Maria Piñana; Rodrigo Vásquez; Tomàs Pumarola |
| EPI_ISL_7156454 | Hospital Ángeles Lomas | Instituto de diagnóstico y Referencia Epidemiologicos (INDRE) | Abril Rodriguez-Maldonado; Ariadna Medina-Benitez; Armando Rojo; Claudia Wong-Arambula; Ernesto Ramirez-Gonzalez; Fernando Gonzalez-Dominguez; Gisela Barrera-Badillo; Irma Lopez-Martinez; Joaquin Quiroz-Mercado; Leonardo Medina Arias; Lucia Hernandez-Rivas; Maribel Gonzalez-Villa; Natividad Cruz-Ortiz; Pilar Escamilla Llano; Raymundo Rodríguez Sandoval; Tatiana Nunez-Garcia; Vanessa Rivero-Arredondo |
| EPI_ISL_7415721, EPI_ISL_7415723, EPI_ISL_7415731, EPI_ISL_7415770, EPI_ISL_7415774, EPI_ISL_7415830 | Houston Methodist Hospital | Houston Methodist Hospital | Ilya J. Finkelstein; James J. Davis; Jessica Cambric; Jimmy Gollihar; Kristina Reppond; Layne Pruitt; Madison N. Shyer; Marcus Nguyen; Matthew Ojeda Saavedra; Paul A. Christensen; Prasanti Yerramilli; Randall J. Olsen; Robert Olson; Ryan Gadd; S. Wesley Long; Sishir Subedi; and James M. Musser |
| EPI_ISL_5858874, EPI_ISL_5858879, EPI_ISL_5858893, EPI_ISL_5858900, EPI_ISL_5858936 | IDIME - Sede Clínica Nueva de Cali | Universidad del Valle | Andres Castillo; Beatriz Parra & Programa Nacional de Caracterización Genómica de SARS-CoV-2; Diana López-Alvarez; Erica M. Aristizabal; Flor Saa; Melissa Solarte; Nelson Rivera Franco |
| EPI_ISL_7451096 | INSA | Instituto Nacional de Saude (INSA) | Borges et al |
| EPI_ISL_6128894, EPI_ISL_6128908, EPI_ISL_6128920, EPI_ISL_6128928, EPI_ISL_6128938, EPI_ISL_6128951, EPI_ISL_6128959, EPI_ISL_6128997, EPI_ISL_6129008, EPI_ISL_6129023, EPI_ISL_6129027, EPI_ISL_6129052, EPI_ISL_6129061, EPI_ISL_6129068, EPI_ISL_6129090, EPI_ISL_6129096, EPI_ISL_6129104, EPI_ISL_6129110, EPI_ISL_6129119, EPI_ISL_6129124, EPI_ISL_6129144, EPI_ISL_6129160, EPI_ISL_6129169, EPI_ISL_6129174, EPI_ISL_6129181, EPI_ISL_6129193, EPI_ISL_6129200, EPI_ISL_6129206, EPI_ISL_6208290, EPI_ISL_6208320, EPI_ISL_6334953, EPI_ISL_6334959, EPI_ISL_6334962 | | | |
| see above | INSPI-CRN DE INFLUENZA Y OTROS VIRUS RESPIRATORIOS | INSPI-CRN DE INFLUENZA Y OTROS VIRUS RESPIRATORIOS | Alfredo Bruno; Carlos Chiluiza; Daniel Ramos; Domenica de Mora; Fernando Llerena; Jimmy Garcés; Lizbeth Patiño; Marcela Mejia; Maria Angelica Becerra; Maritza Olmedo; Michelle Páez; Ruben Armas |
| EPI_ISL_3274403 | INSPI-CRN DE INFLUENZA Y OTROS VIRUS RESPIRATORIOS | NIC-INSPI | Alfredo Bruno; Domenica de Mora.; Jimmy Garcés; Johanna Laines; Lizbeth Patiño; Manuel Gonzalez; Maritza Olmedo; Michelle Páez |
| EPI_ISL_5796708, EPI_ISL_6265022, EPI_ISL_6265112, EPI_ISL_6265164, EPI_ISL_6815686, EPI_ISL_6815692, EPI_ISL_6815739, EPI_ISL_6815747, EPI_ISL_6815750, EPI_ISL_6815756, EPI_ISL_6815835 | | | |
| see above | IRCCS San Gallicano Dermatological Institute | IRCCS Regina Elena National Cancer Institute | Aldo Morrone; Alessia Lauretti; Alice Massacci; Andrea Cazzani; Antonio Federico; Carla Mottini; Eleonora Sperandio; Elisabetta Trento; Fabrizio Ensoli; Francesca De Nicola; Francesca Maione; Francisco Obregon; Frauke Goeman; Fulvia Pimpinelli; Gennaro Ciliberto; Giovanni Blandino; Giulia Orlandi; Ludovica Ciuffreda; Martina Betti; Martina Diano; Matteo Pallocca; Maurizio Fanciulli; Sabrina Strano; Sara Donzelli; Serena Salvo |
| EPI_ISL_6102799 | Infinity Biologix | Centers for Disease Control and Prevention Division of Viral Diseases, Pathogen Discovery | Benjamin Rambo-Martin; Chirayu Goswami; Christian Bixby; Christopher Gulvick; Clinton Paden; Dakota Howard; Dhwani Batra; Duncan MacCannell; Erisa Sula; Jason Caravas; Jonathan Schultz; Kristine Lacek; Matthew Schmerer; Peter Cook; Robin Grimwood; Russ Hager; Scott Sammons; Shatavia Morrison; Tymeckia Kendall; Victoria Caban Figueroa; Yihe Wang; Yvette Unoarumhi |
| EPI_ISL_6944125 | KEMRI-Wellcome Trust Research Programme,Kilifi | KEMRI-Wellcome Trust Research Programme,Kilifi | Agoti C.; D.J Nokes; Githinji G.; Lambisia A. Makori T.; Mburu M.W; Mohamed K.S.; Morobe J.; Ndwiga L.; Ochola I; Ongera E.; de Laurent Z. |
| EPI_ISL_6794907, EPI_ISL_6989250, | KU Leuven, Rega Institute, Clinical and | KU Leuven, Rega Institute, Clinical and Epidemiological Virology | Bert Vanmechelen; Casper Geenen; Emmanuel André; Guy Baele; Joan Marti-Carerras; Joren; Lize Cuypers; Piet Maes; Raymenants; Sarah Gorissen; Simon Dellicour; Tony Wawina-Bokalanga |

| | | | |
|---|---|---|---|
| EPI_ISL_7413964 | Epidemiological Virology | | |
| EPI_ISL_7196031 | Karolinska University Hospital Huddinge | Karolinska University Hospital | Annelie Bjerkner; Isak Sylvin; Jan Albert; Karolina Ininbergs; Lina Guerra Blomqvist; Lynda Eneh; Martin Ekman; Martina Wahlund; Robert Dyrdak; Sandra Broddesson; Tanja Normark; Tobias Allander; Valtteri Wirta; Zhibing Yun |
| EPI_ISL_6324708, EPI_ISL_6377103 | Karolinska University Hospital Solna | Karolinska University Hospital | Annelie Bjerkner; Isak Sylvin; Jan Albert; Karolina Ininbergs; Lina Guerra Blomqvist; Lynda Eneh; Martin Ekman; Martina Wahlund; Robert Dyrdak; Sandra Broddesson; Tanja Normark; Tobias Allander; Valtteri Wirta; Zhibing Yun |
| EPI_ISL_7611232 | LABORATOIRE BIOALLIANCE | CNR Virus des Infections Respiratoires - France SUD | Antonin Bal; Bruno Lina; Bruno Simon; Gregory Destras; Gwendolyne Burfin; Hadrien Regue; Laurence Josset; Martine Valette; Quentin Semanas |
| EPI_ISL_5914084 | LABORATOIRE BIOESTEREL | CNR Virus des Infections Respiratoires - France SUD | Antonin Bal; Bruno Lina; Bruno Simon; Gregory Destras; Gwendolyne Burfin; Hadrien Regue; Laurence Josset; Martine Valette; Quentin Semanas |
| EPI_ISL_7611375 | LABORATOIRE BIOFUSION | CNR Virus des Infections Respiratoires - France SUD | Antonin Bal; Bruno Lina; Bruno Simon; Gregory Destras; Gwendolyne Burfin; Hadrien Regue; Laurence Josset; Martine Valette; Quentin Semanas |
| EPI_ISL_7610788, EPI_ISL_7610793 | LABORATOIRE MAYMAT | CNR Virus des Infections Respiratoires - France SUD | Antonin Bal; Bruno Lina; Bruno Simon; Gregory Destras; Gwendolyne Burfin; Hadrien Regue; Laurence Josset; Martine Valette; Quentin Semanas |
| EPI_ISL_6602302 | LABORATOIRE SYNLAB / LABIO MAS | CNR Virus des Infections Respiratoires - France SUD | Antonin Bal; Bruno Lina; Bruno Simon; Gregory Destras; Gwendolyne Burfin; Hadrien Regue; Laurence Josset; Martine Valette; Quentin Semanas |
| EPI_ISL_6721424, EPI_ISL_6721435, EPI_ISL_6721437, EPI_ISL_6721443, EPI_ISL_6721452, EPI_ISL_6721462, EPI_ISL_6721475, EPI_ISL_6721482, EPI_ISL_6721490, EPI_ISL_6721512, EPI_ISL_6721520, EPI_ISL_6721529, EPI_ISL_6721535, EPI_ISL_6721791, EPI_ISL_6721799, EPI_ISL_6721805, EPI_ISL_6721809 | | | |
| see above | LACEN - Laboratório Central de Saúde Pública de Pernambuco | Evandro Chagas Institute | Delana Andreza Melo Bezerra; Dielle Monteiro Teixeira; Jedson Ferreira Cardoso; Jessylene de Almeida Ferreira; Kenny da Costa Pinheiro; Luana Soares Barbagelata; Luana da Silva Soares; Mirleide Cordeiro dos Santos; Patrícia dos Santos Lobo; Rayssa Layna da Silva Bedran; Sandro Patroca da Silva |
| EPI_ISL_6721266, EPI_ISL_6721279, EPI_ISL_6721282, EPI_ISL_6721290, EPI_ISL_6721300, EPI_ISL_6721306, EPI_ISL_6721313, EPI_ISL_6721768 | | | |
| see above | LACEN - Laboratório Central de Saúde Pública de Roraima | Evandro Chagas Institute | Delana Andreza Melo Bezerra; Dielle Monteiro Teixeira; Jedson Ferreira Cardoso; Jessylene de Almeida Ferreira; Kenny da Costa Pinheiro; Luana Soares Barbagelata; Luana da Silva Soares; Mirleide Cordeiro dos Santos; Patrícia dos Santos Lobo; Rayssa Layna da Silva Bedran; Sandro Patroca da Silva |
| EPI_ISL_6721542, EPI_ISL_6721679, EPI_ISL_6721705, EPI_ISL_6721707, EPI_ISL_6721710 | LACEN - Laboratório Central de Saúde Pública do Amazonas | Evandro Chagas Institute | Delana Andreza Melo Bezerra; Dielle Monteiro Teixeira; Jedson Ferreira Cardoso; Jessylene de Almeida Ferreira; Kenny da Costa Pinheiro; Luana Soares Barbagelata; Luana da Silva Soares; Mirleide Cordeiro dos Santos; Patrícia dos Santos Lobo; Rayssa Layna da Silva Bedran; Sandro Patroca da Silva |
| EPI_ISL_6721559, EPI_ISL_6721565, EPI_ISL_6721573, EPI_ISL_6721582, EPI_ISL_6721587, EPI_ISL_6721593, EPI_ISL_6721597, EPI_ISL_6721604, EPI_ISL_6721608, EPI_ISL_6721612, EPI_ISL_6721618, EPI_ISL_6721631, EPI_ISL_6721634, EPI_ISL_6721639, EPI_ISL_6721648, EPI_ISL_6721668, EPI_ISL_6721675, EPI_ISL_6721834 | | | |
| see above | LACEN - Laboratório Central de Saúde Pública do Ceara | Evandro Chagas Institute | Delana Andreza Melo Bezerra; Dielle Monteiro Teixeira; Jedson Ferreira Cardoso; Jessylene de Almeida Ferreira; Kenny da Costa Pinheiro; Luana Soares Barbagelata; Luana da Silva Soares; Mirleide Cordeiro dos Santos; Patrícia dos Santos Lobo; Rayssa Layna da Silva Bedran; Sandro Patroca da Silva |
| EPI_ISL_6721332, EPI_ISL_6721336, EPI_ISL_6721339, EPI_ISL_6721346, EPI_ISL_6721355, EPI_ISL_6721359, EPI_ISL_6721368, EPI_ISL_6721376, EPI_ISL_6721385, EPI_ISL_6721390, EPI_ISL_6721396, EPI_ISL_6721401, EPI_ISL_6721419, EPI_ISL_6721776, EPI_ISL_6721784 | | | |
| see above | LACEN - Laboratório Central de Saúde Pública do Rio Grande do Norte | Evandro Chagas Institute | Delana Andreza Melo Bezerra; Dielle Monteiro Teixeira; Jedson Ferreira Cardoso; Jessylene de Almeida Ferreira; Kenny da Costa Pinheiro; Luana Soares Barbagelata; Luana da Silva Soares; Mirleide Cordeiro dos Santos; Patrícia dos Santos Lobo; Rayssa Layna da Silva Bedran; Sandro Patroca da Silva |
| EPI_ISL_6704864, EPI_ISL_6704867, EPI_ISL_6704870, EPI_ISL_6704874 | LANCET LABORATORY | National Institute for Communicable Diseases of the National Health Laboratory Service | Amoako DG; Bhiman JN; Everatt J; Ismail A; Mahlangu B; Mnguni A; Mohale T; Ntuli N; Scheepers C; Wolter N |
| EPI_ISL_6901960, EPI_ISL_6901961, EPI_ISL_7473154 | LATE - Laboratório de Técnicas Especiais - Hospital Israelita Albert Einstein | LATE - Laboratório de Técnicas Especiais - Hospital Israelita Albert Einstein | Alexandre Hideaki Takara; Ana Paula Moreira Salles; Anelisie da Silva Santos; Deyvid Amgarten; Erick Gustavo Dorlass; Fernanda de Mello Malta; João Renato Rebello Pinho; Luiz Vicente Rizzo; Marcio Anunciacao Menezes; Pedro Henrique Sebe Rodrigues; Raquel Riyuzo |
| EPI_ISL_6316493, EPI_ISL_6316495 | LBM Porte de la Chapelle | CERBA HealthCare | Bénédicte Roquebert; Laura Verdurme; Sabine Trombert; Stéphanie Haim-Boukobza |
| EPI_ISL_3460003 | LESP Tabasco | Instituto de Diagnostico y Referencia Epidemiologicos (INDRE) | Abril Rodriguez-Maldonado; Ariadna Medina-Benitez; Claudia Wong-Arambula; Ernesto Ramirez-Gonzalez.; Gisela Barrera-Badillo; Irma Lopez-Martinez; Joaquin Quiroz-Mercado; Lucia Hernandez-Rivas; Maribel Gonzalez-Villa; Natividad Cruz-Ortiz; Sergio Rangel-Guerrero; Tatiana Nunez-Garcia; Vanessa Rivero-Arredondo |
| EPI_ISL_5923350, EPI_ISL_5923355, EPI_ISL_5923358 | LIME | Laboratorio Departamental de Salud Publica de Antioquia | Ana Victoria Valencia Duarte; Cristian Arbey Velarde Hoyos; Gloria Isabel Escobar; Idabely Betancur Ortiz; Juan P. Hernandez-Ortiz; Juan Pablo Isaza Agudelo; Maria Stella López |
| EPI_ISL_5887476, EPI_ISL_5887478, EPI_ISL_5887479 | Lab. Hospital clinico Universidad de Chile | "Facultad de Ciencias de la Vida, UNAB" | "Claudio Meneses; Ariel Orellana"; Claudio Olmos; Daniel Leon; Dayan Sanhueza; Eduardo Castro; Gonzalo Campaña; Macarena Bastias; Paola Pidal; Ricardo Yusta; Sebastian Wolter; Susana Saez; Víctor Monreal; Waldo Diaz |
| EPI_ISL_6116520, EPI_ISL_6315910, EPI_ISL_6424828, EPI_ISL_6699342, EPI_ISL_6699432, EPI_ISL_6699466, EPI_ISL_6885031, EPI_ISL_6886973, EPI_ISL_6887009, EPI_ISL_7218603, EPI_ISL_7218764, EPI_ISL_7268416, EPI_ISL_7460273, EPI_ISL_7460312 | | | |
| see above | Labo Analyses Med | National Reference Center for Viruses of Respiratory Infections, Institut Pasteur, Paris | Angela Brisebarre; Brieuc Gestin; Camille Capel; Caroline MALDERET; Christophe Malabat; Claire Felloni; Corinne Maufrais; Domitille LEMAN (59200); Etienne Simon-Lorière; Frédéric Lemoine; Julien Fumey; Karine Michez; Louise Lefrançois; Marion Barbet; Maud Vanpeene; Méline Bizard; Slim El Khiari; Slim El-Khiari; Sylvie Behillil; Sylvie Van der Werf; Thierry Guffond; Vincent Enouf |
| EPI_ISL_5655471, EPI_ISL_5655472, EPI_ISL_5655473, EPI_ISL_5655474, EPI_ISL_5655475 | Laboratoire BIORANCE | CHU Pontchaillou | DE TAYRAC Marie; DENOUAL Florent; ETCHEVERRY Amandine; FEBREAU Christine; GALIBERT Marie Dominique; GROLHIER Claire; JAGLINE Steven; PRONIER Charlotte; QUENET Benjamin; SASSI Mohamed; THIBAULT Vincent |
| EPI_ISL_7550845, EPI_ISL_7550846, EPI_ISL_7550847, EPI_ISL_7550848, EPI_ISL_7550849, EPI_ISL_7550852, EPI_ISL_7550853, EPI_ISL_7550854, EPI_ISL_7550855, EPI_ISL_7550856, EPI_ISL_7550857, EPI_ISL_7550858, EPI_ISL_7550860, EPI_ISL_7550862, EPI_ISL_7550864, EPI_ISL_7550866, EPI_ISL_7550867, EPI_ISL_7550868, EPI_ISL_7550869, EPI_ISL_7550870, EPI_ISL_7550871, EPI_ISL_7550872, EPI_ISL_7550873, EPI_ISL_7550874, EPI_ISL_7550875, EPI_ISL_7550876, EPI_ISL_7550877, EPI_ISL_7550878, EPI_ISL_7550879, EPI_ISL_7550880, EPI_ISL_7550881, EPI_ISL_7550882, EPI_ISL_7550883, EPI_ISL_7550884, EPI_ISL_7550885, EPI_ISL_7550886, EPI_ISL_7550888, EPI_ISL_7550892, EPI_ISL_7550893, EPI_ISL_7550894, EPI_ISL_7550896, EPI_ISL_7550897, EPI_ISL_7550899, EPI_ISL_7550901, EPI_ISL_7550902, EPI_ISL_7550903, EPI_ISL_7550904, EPI_ISL_7550905, EPI_ISL_7550906, EPI_ISL_7550908, EPI_ISL_7550909, EPI_ISL_7550910, EPI_ISL_7550913, EPI_ISL_7550914, EPI_ISL_7550915, EPI_ISL_7550917, EPI_ISL_7550918, EPI_ISL_7550919, EPI_ISL_7550920, EPI_ISL_7550921, EPI_ISL_7550922, EPI_ISL_7550923, EPI_ISL_7550924, EPI_ISL_7550925, EPI_ISL_7550926, EPI_ISL_7550927, EPI_ISL_7550929, EPI_ISL_7550931, EPI_ISL_7550937, EPI_ISL_7550939, EPI_ISL_7550940, EPI_ISL_7550942, EPI_ISL_7550943, EPI_ISL_7550944, EPI_ISL_7550946, EPI_ISL_7550947, EPI_ISL_7550948, EPI_ISL_7550949, EPI_ISL_7550950, EPI_ISL_7550951, EPI_ISL_7550952, EPI_ISL_7550954, EPI_ISL_7550955, EPI_ISL_7550956, EPI_ISL_7550957, EPI_ISL_7550958, EPI_ISL_7550964, EPI_ISL_7550967, EPI_ISL_7550968, EPI_ISL_7550969, EPI_ISL_7550970, EPI_ISL_7550971, EPI_ISL_7550972, EPI_ISL_7550974, EPI_ISL_7550977, EPI_ISL_7550978, EPI_ISL_7550979, EPI_ISL_7550981, EPI_ISL_7550986, EPI_ISL_7550987, EPI_ISL_7550989, EPI_ISL_7550991, EPI_ISL_7550993, EPI_ISL_7550995, EPI_ISL_7550996, EPI_ISL_7550997, EPI_ISL_7550998, EPI_ISL_7551000, EPI_ISL_7551001, EPI_ISL_7551002 | | | |
| see above | Laboratoire de santé publique du Québec | Laboratoire de santé publique du Québec | Guillaume Bourque; Ioannis Ragoussis; Jesse Shapiro; Mark Lathrop and Judith Fafard on behalf of the CoVSeQ research group; Sandrine Moreira |
| EPI_ISL_2271701 | Laboratorio Central, Ministerio de Salud Cordoba | Instituto de Patologia Vegetal (CIAP-INTA) on behalf of 'Proyecto Argentino Interinstitucional de genomica de SARS-CoV-2' (PAIS Consortium) | Barbas, G.; Castro, G.; Debat, HJ.; FD; Fernandez; M; M.B.; Marquez, N.; Pisano; Re, V. |
| EPI_ISL_4503176 | Laboratorio de Biología Molecular de la Universidad Nacional de San Agustín de Arequipa | Laboratorio de Recursos Genéticos y Genética Molecular de la Universidad Nacional de San Agustín de Arequipa | Alejandra Dávila; Guillermo Salvatierra; Jorge Ballón; Kasandra Ascuña; Pablo Tsukayama; Patrick Fernández; Renzo Salazar; Rosario Valderrama |
| EPI_ISL_4405149 | Laboratorio de Biología molecular del Hospital General de Agudos Dr. Carlos G. Durand | Área de Secuenciación del Laboratorio de Virología del Hospital de Niños Dr. Ricardo Gutierrez on behalf of 'Proyecto Argentino Interinstitucional de genomica de SARS-CoV-2' (PAIS Consortium) | A; Acuña; Anzorena; B; C; Carrón; Castro; Cañellas; Chiussi; Colina; Costa; D; Dahinten; Dima; Domínguez; E; Elsegood; F; Frisone; G; Gatica; Goya; Irrazábal; Jurado; L; LE; Leivas; Loayza; Lusso; M; MB; MF; MI; MS; Marchissio; Marina; Matillas; N; Nabaes Jodar; Narduzzi; Natale; Notaristéfano; R; Rivarola; Rodríguez Cardozo; Rodríguez Saá; Rozo; S; Theaux; Valinotto; Viegas, M.; W; Warszatska; Y; Yaunguzian |
| EPI_ISL_3655695, EPI_ISL_5495023, EPI_ISL_5495032, EPI_ISL_5495041, EPI_ISL_5495053, EPI_ISL_5495060, EPI_ISL_5495064, EPI_ISL_5495075, EPI_ISL_5495093, EPI_ISL_5495101, EPI_ISL_5495128, EPI_ISL_5495147, EPI_ISL_5495151, EPI_ISL_5495158, EPI_ISL_5495167, EPI_ISL_5495174, EPI_ISL_5495184, EPI_ISL_5495194, EPI_ISL_5495218, EPI_ISL_5495233, EPI_ISL_5495237, EPI_ISL_5495240, EPI_ISL_5495281, EPI_ISL_5495286, EPI_ISL_5495300, EPI_ISL_5495309, EPI_ISL_5495336, EPI_ISL_5495360, EPI_ISL_5495377, EPI_ISL_5495393, EPI_ISL_5495397, EPI_ISL_5495403, EPI_ISL_5495407, EPI_ISL_5495413, EPI_ISL_5495419, EPI_ISL_5495428, EPI_ISL_5495433, EPI_ISL_5495467, EPI_ISL_5495479, EPI_ISL_5495487, EPI_ISL_5495491, EPI_ISL_5495504, EPI_ISL_5495510, EPI_ISL_5495518, EPI_ISL_5495533, EPI_ISL_5495548, EPI_ISL_5495550, EPI_ISL_5495553, EPI_ISL_5495567, EPI_ISL_5495573, EPI_ISL_5495578, EPI_ISL_5764291, EPI_ISL_5764461, EPI_ISL_5764522 | | | |
| see above | Laboratorio de Referencia Nacional de Virus Respiratorios. Centro Nacional de Salud Publica. Instituto Nacional de Salud Peru. | Laboratorio de Referencia Nacional de Virus Respiratorios. Centro Nacional de Salud Publica. Instituto Nacional de Salud Peru. | Carlos Padilla Rojas; Henri Bailon Calderon; Iris Silva Molina; Joseph Huayra Niquen; Lely Solari Zerpa; Luis Barcena Flores; Marco Galarza Perez; Nancy Rojas Serrano; Nieves Sevilla Castañeda; Omar Caceres Rey; Orson Mestanza Millones; Princesa Medrano Alhuay; Priscila Lope Pari; Sandra Morales Ruiz; Sara Gordillo Vilchez; Steve Acedo Lazo; Veronica Hurtado Vela; Victor Jimenez Vasquez; Wendy Lizarraga Olivares |
| EPI_ISL_2921332, EPI_ISL_2921334, EPI_ISL_2921361, EPI_ISL_2921362, EPI_ISL_2921363, EPI_ISL_2921365, EPI_ISL_2921368, EPI_ISL_2921369, EPI_ISL_2921371 | | | |

| | | | |
|---|---|---|---|
| see above | Laboratorio de Referencial Nacional de Virus Respiratorios | Laboratorio de Referencial Nacional de Virus Respiratorios | Carlos Padilla Rojas; Henri Bailon Calderon; Iris Silva Molina; Joseph Huayra Niquen; Lely Solari Zerpa; Luis Barcena Flores; Marco Galarza Perez; Nancy Rojas Serrano; Omar Caceres Rey; Orson Mestanza Millones; Priscila Lope Pari; Sandra Morales Ruiz; Steve Acedo Lazo; Veronica Hurtado Vela |
| EPI_ISL_3275319 | Laboratorio de Vigilancia en Salud Pública el Salvador | Genomics and Proteomics Departament, Gorgas Memorial Institute For Health Studies | Alexander Martinez; Ambar Moreno; Claudia Díaz; Claudia Gonzalez; Denis G Jovel A; Gustavo M Ramirez; Jessica Gondola; Leyda Abrego; Marlene Castillo; Oris Chavarria; Ruth C Vasquez C; Sandra Paola Paz |
| EPI_ISL_3017570 | Laboratory Corporation of America | Centers for Disease Control and Prevention Division of Viral Diseases, Pathogen Discovery | Adrian Paskey; Amanda Douglas; Amanda Suchanek; Andrea Throop; Ayla Burns; Benjamin Rambo-Martin; Bobbi Croy; Brian Krueger; Brian Norvell; Christopher Gulvick; Christos Petropoulos; Clinton R. Paden; Craig Lukasik; Dakota Howard; Darlene Wagner; Debbie Boles; Dhwani Batra; Duncan MacCannell; Eyad Almasri; Goran Stevovic; Howard Engler; Hrushikesh Deshmukh; Jake Humphrey; Jana Schroth; Jason Caravas; Joe Voshell; John Pruitt; Jonathan Meltzer; Jonathan Williams; Kara Moser; Kimberly Wagner; Lax Iyer; Lisa Pfefferle; Lyndon Tilson; Manoj Jain; Marcia Eisenberg; Mary Ann Cristobal; Mary Williamson; Matthew Robinson; Matthew Schmerer; Michael Levandoski; Mike Sapeta; Mindy Nye; Minoo Agarwal; Mohan Kolli; Nuthawin Charoensri; Oren Cohen; Peter W. Cook; Prashant Gupta; Qian Zeng; Rama Ghatti; Scott Parker; Scott Ryan; Scott Sammons; Shatavia Morrison; Stanley Letovsky; Steven Ragan; Suresh Babu Selvaraju; Susan Countryman; Susan Hicks; Suzanne Dale; Thomas Urban; Tim Kuphal; Tricia Zwiefelhofer; Vincent Drouillon; Yvette Unoarumhi |
| EPI_ISL_7220176 | Laboratory of Clinical Virology Heraklion Crete | Laboratory of Clinical Virology Heraklion Crete | Alexandros Zafiropoulos; George Sourvinos |
| EPI_ISL_6774037 | Lahad Datu Hospital | Institute for Medical Research, Infectious Disease Research Centre, National Institutes of Health, Ministry of Health Malaysia | Ahmad FA; Ahmad Fazilah NA; Anasir MI; Azizan MA; Kamel K; Mohamad Sukri MZ; Mohd Zawawi Z; Norhisham SN; Ramly N; Robert F; Rosli NR; Suppiah J; Thayan R |
| EPI_ISL_6914961 | Lancet | NHLS/UCT | Arash Iranzadeh; Bruna Galvao; Carolyn Williamson; Deelan Doolabh; Diana Hardie; Gert Marais; Innocent Mudau; Lynn Tyers; Marvin Hsiao; Rageema Joseph; Stephen Korsman |
| EPI_ISL_6913991, EPI_ISL_6914000 | Lancet Laboratories | National Institute for Communicable Diseases of the National Health Laboratory Service | Amoako DG; Bhiman JN; Everatt J; Ismail A; Mahlangu B; Mnguni A; Mohale T; Ntuli N; Scheepers C; Wolter N |
| EPI_ISL_7353252 | Lighthouse Lab in Glasgow | Wellcome Sanger Institute for the COVID-19 Genomics UK (COG-UK) Consortium | Anna Dominiczak and Alex Alderton; Carol Clugston; Cordelia Langford; David Gray; David K. Jackson; Dominic Kwiatkowski; Ewan Harrison; Harper VanSteenhouse; Ian Johnston; Jeffrey Barrett; John Sillitoe on behalf of the Wellcome Sanger Institute COVID-19 Surveillance Team; Roberto Amato; Sonia Goncalves; Yumi Kasai |
| EPI_ISL_7181977, EPI_ISL_7586776 | Lighthouse Lab in Milton Keynes | Wellcome Sanger Institute for the COVID-19 Genomics UK (COG-UK) Consortium | Cordelia Langford; David K. Jackson; Dominic Kwiatkowski; Ewan Harrison; Ian Johnston; Jeffrey Barrett; John Sillitoe on behalf of the Wellcome Sanger Institute COVID-19 Surveillance Team; Roberto Amato; Sonia Goncalves; The Lighthouse Lab in Milton Keynes and Alex Alderton |
| EPI_ISL_7413422, EPI_ISL_7413427, EPI_ISL_7413439 | MB-Cadham Provincial laboratory | National Microbiology Laboratory (NML) | Anna Majer; Anneliese Landgraff; CanCOGeN's metadata curation team; Darian Hole; David Alexander; Elsie Grudeski; Gary Van Domselaar; Grace Seo; Jared Bullard; Jennifer Tanner; Kerry Dust; Kirsten Biggar; Madison Chapel; Morag Graham; Natalie Knox; Nathalie Bastien; Paul Van Caeseele; Philip Mabon; Public Health Agency of Canada CanCOGeN team; Rhiannon Huzarewich; Russell Mandes; Shari Tyson; Timothy Booth; Yan Li |
| EPI_ISL_6492635, EPI_ISL_6492796, EPI_ISL_6492832 | MSHS Clinical Microbiology Laboratories | MSHS Pathogen Surveillance Program | Adolfo García-Sastre; Adriana van de Guchte; Ajay Obla; Alberto Paniz-Mondolfi; Ana S. Gonzalez-Reiche; Angela Amoako; Ashley Salimbangon; Betsaida Salom Melo; Bremy Alburquerque; Brianne Ciferri; Charles Gleason; Daniel Floda; Deena R. Altman; Denise Jurczyszak; Emilia Mia Sordillo; Gintaras Deikus; Giulio Kleiner; Gopi Patel; Hala Alshammary; Harm van Bakel; Irina Oussenko; Jayeeta Dutta; Juan Soto; Julia Matthews; Katherine Beach; Kathryn Twyman; Kayla Russo; Komal Srivastava; Levy Sominsky; Mahmoud Awawda; Marta Luksza; Matthew M. Hernandez; Melissa Gitman; Michael D. Nowak; Mitchell J. Sullivan; Nancy Francoeur; Robert Sebra; Sarah Schaefer; Shelcie Fabre; Shwetha Hara Sridhar; Viviana Simon; Ying-Chih Wang; Zain Khalil; Zenab Khan |
| EPI_ISL_6760707, EPI_ISL_6760905, EPI_ISL_6760973, EPI_ISL_6764745, EPI_ISL_6768069 | Macha Research Trust | Macha Research Trust | Gill CJ; Kalonda A; Katowa B; Kwenda G; Matoba J; Mubemba B; Muleya W; Mupila Z; Mwananyanda L; Simulundu E |
| EPI_ISL_6963049, EPI_ISL_6963052, EPI_ISL_6963118, EPI_ISL_6963122, EPI_ISL_6963124 | Malawi Liverpool Wellcome Trust Clinical Research Program | Malawi Liverpool Wellcome Trust Clinical Research Program | Ben Morton; Catherine Anscombe; Kondwani Jambo; Philip Ashton; Sam Lissauer |
| EPI_ISL_3435363 | Maryland Genomics, Institute for Genome Sciences, University of Maryland School of Medicine | Maryland Genomics, Institute for Genome Sciences, University of Maryland School of Medicine | Claire M; Fraser; Hazen; Holly; Humphrys; Ivette; Jacques; Jonathan; Kranthi; Lim; Lisa D; Luke J; Mike; Ott; Ravel; Roussey; Sadzewicz; Sandra; Santana-Cruz; Tallon; Tracy; Vavikolanu |
| EPI_ISL_7425654, EPI_ISL_7443804, EPI_ISL_7443805 | Medizinische Laboratorien Düsseldorf | Robert Koch Institute | |
| EPI_ISL_6854347 | Microbiologia e Virologia Cotugno | Microbiologia e Virologia Cotugno | Antonio Canonico; Antonio Fascione; Claudia Tiberio; Enza Mallardo; Francesco Nappo; Giovanni D'Auria; Giuseppe di Gennaro; Ilaria Cavallaro; Luigi Atripaldi |
| EPI_ISL_6575650, EPI_ISL_6575660, EPI_ISL_6575662, EPI_ISL_6575665, EPI_ISL_6623843, EPI_ISL_6631667, EPI_ISL_6639752, EPI_ISL_6641078, EPI_ISL_6698641, EPI_ISL_6878318, EPI_ISL_6914394 | | | |
| see above | Molecular Virology section, department of infection & immunity, KFSHRC | Molecular Virology section, department of infection & immunity, KFSHRC | Ahmed A. Al-Qahtani; Basma M. Alahideb; Fatimah S. Alhamlan; Feda A. Alsuwairi; Maha Al-Abdulkarim; Maysoon S. Mutabagani; Mohamed S. Asiri; Reem S. Almaghrabi Madain S. Alsanea; Sahar I. Althawadi; Sara A. Altamimi |
| EPI_ISL_4772303 | NAMRU-6 | NAMRU-6 | Cristhopher Cruz; Eugenio Abente; Francis Chuquirachi Panduro; Gilda Troncos; Greg Rice; Marita Silva; Paul Graf.; Sonia Ampuero; Victoria Espejo; Yeny Tinoco |
| EPI_ISL_7545672 | NHLS Livingstone Laboratory | CERI, Centre for Epidemic Response and Innvoation, Stellenbosch University and KRISP, KZN Research Innovation and Sequencing Platform, UKZN. | Arisha Maharaj; Giandhari J; Naidoo Y; Oluwakemi Laguda-Akingba and Nokukhanya Mdlalose; Pillay S; Ramphal U; Ramphal Y; San JE; Tegally H; Tshiabuila D; Wilkinson E; de Oliveira T |
| EPI_ISL_6506316, EPI_ISL_6506321, EPI_ISL_6506329, EPI_ISL_6506366, EPI_ISL_6506405, EPI_ISL_6506424, EPI_ISL_6506435, EPI_ISL_6506446, EPI_ISL_6506496, EPI_ISL_6506549, EPI_ISL_6506569, EPI_ISL_6506573, EPI_ISL_6506592, EPI_ISL_6506600, EPI_ISL_6506610 | | | |
| see above | Namibia Institute of Pathology LTD | CERI, Centre for Epidemic Response and Innvoation, Stellenbosch University and KRISP, KZN Research Innovation and Sequencing Platform, UKZN. | Andreas Shiningavamwe; Giandhari Jennifer; Iyaloo Constantinus; Naidoo Yeshnee; Nathalia Garus-Oas; Ndahafa Frans; Ndumbu Pentikainen; Pillay Sureshnee; San James; Tegally Houriiyah; Tshiabuila Derek; Wilkinson Eduan; Yajna Ramphal; de Oliveira Tulio |
| EPI_ISL_7310648, EPI_ISL_7310685, EPI_ISL_7310733 | National Health Laboratory Service, Kwazulu-Natal, South Africa | CERI, Centre for Epidemic Response and Innvoation, Stellenbosch University and KRISP, KZN Research Innovation and Sequencing Platform, UKZN. | Arisha Maharaj; Giandhari J; Moir M; Naidoo Y; Nokukhanya Mdlalose; Pillay S; Ramphal U; Ramphal Y; San JE; Tegally H; Tshiabuila D; Wilkinson E; de Oliveira T |
| EPI_ISL_6699742, EPI_ISL_6699764 | National Health Laboratory Services, Virology, Charlotte Maxeke Johannesburg hospital, Parktown, Johannesburg, Gauteng | CERI, Centre for Epidemic Response and Innvoation, Stellenbosch University and KRISP, KZN Research Innovation and Sequencing Platform, UKZN. | Amoaka D; Arisha Maharaj; Avani Bharuthram; Bester P; Bhiman J; Engelbrecht S; Everatt J; Florette Treurnicht; Goedhals D; Hardie D; Hsiao M; Iranzadeh A; Kathleen Subramoney; Lessells R; Makatini Z; Maponga T; Mdlalose N; Mlisana K; Moir M; NGS-SA (Scheepers C; Naidoo Y; Nkhensani Mtileni; Nyaga M) Giandhari J; Oluwakemi M; Pillay S; Preiser W; Ramphal U; Ramphal Y; San JE; Tegally H; Tshiabuila D; Venter M; Wilkinson E; Williamson C; de Oliveira T; von Gottberg A |
| EPI_ISL_6939036 | National Influenza Centre | National Influenza Centre | ; Benjamiin B. Lindsey; Benjamin H. Foulkes; Bless Seyram Agbenyo; Bright Adu; Ernest Asiedu; Franklin Asiedu-Bekoe; Hilda Opoku Frempong; Ivy A. Asante; Joseph Oliver-Commey; Joyce Appiah-Kubi; Keren Okyerebea Attiku; Linda Boatemaa; Lorreta Kwasah; Mathew D. Parker; Michael Marks; Mildred Adusei-Poku; Quaneeta Mohktar; Sharon Hsu; Thushan I de Silva; William K. Ampofo |
| EPI_ISL_7418017 | National Institute of Infectious Diseases | National Institute of Infectious Diseases | Harutaka Katano; Ken Maeda; Kentaro Itokawa; Makoto Kuroda; Shuetsu Fukushi; Shun Iida; Tadaki Suzuki; Tsuyoshi Sekizuka; Yudai Kuroda |
| EPI_ISL_7137310, EPI_ISL_7195620 | National Public Health Laboratory, National Centre for Infectious Diseases | National Public Health Laboratory, National Centre for Infectious Diseases | Benny Yeo Ken Yee; Constance Chen; Dennis Loy Song Qi; Dimitar Kenanov; Grace Ngan Jie Yin; Katherine Ching Zi Yan; Kwan Ki Ko; Lin Cui; Mak Tze Minn; Niranjan Nagarajan; Raymond Tzer Pin Lin; Royce Ang; Samuel Loo; Sebastian Maurer Stroh; Suphavilai Chayaporn; Zhenyang Zhou |
| EPI_ISL_7139725 | National Reference Laboratory, Laboratories Department, Cotonou, Benin | Noguchi Memorial Institute for Medical Research, University of Ghana, Legon, Ghana | Ange D. Dossou; Anges W. Yadouleton; Benjamin I.B. Hounkpatin; Bright Adu; Hilda Opoku Frempong; Joyce Appiah-Kubi; Keren O. Attiku; Quaneeta Mohktar; Rene K. Keke |
| EPI_ISL_7570871, EPI_ISL_7570872, EPI_ISL_7570874, EPI_ISL_7570875, EPI_ISL_7570876, EPI_ISL_7570878, EPI_ISL_7570881, EPI_ISL_7570882, EPI_ISL_7570883, EPI_ISL_7570884, EPI_ISL_7570887, EPI_ISL_7570889, EPI_ISL_7570890, EPI_ISL_7570891, EPI_ISL_7570893, EPI_ISL_7570894, EPI_ISL_7570895, EPI_ISL_7570896, EPI_ISL_7570897, EPI_ISL_7570900, EPI_ISL_7570901, EPI_ISL_7570904, EPI_ISL_7570905, EPI_ISL_7570907, EPI_ISL_7570908, EPI_ISL_7570909, EPI_ISL_7570910, EPI_ISL_7570911, EPI_ISL_7570912, EPI_ISL_7570917, EPI_ISL_7570918, EPI_ISL_7570920, EPI_ISL_7570921, EPI_ISL_7570922, EPI_ISL_7570923, EPI_ISL_7570925, EPI_ISL_7570926, EPI_ISL_7570930, EPI_ISL_7570931, EPI_ISL_7570932, EPI_ISL_7570933, EPI_ISL_7570935, EPI_ISL_7570936, EPI_ISL_7570937, EPI_ISL_7570938, EPI_ISL_7570939, EPI_ISL_7570941, EPI_ISL_7570942, EPI_ISL_7570943, EPI_ISL_7570944, EPI_ISL_7570947, EPI_ISL_7570950, EPI_ISL_7570951, EPI_ISL_7570952, EPI_ISL_7570953, EPI_ISL_7570954, EPI_ISL_7570959, EPI_ISL_7570960, EPI_ISL_7570961, EPI_ISL_7570963, EPI_ISL_7570965, EPI_ISL_7570967, EPI_ISL_7570968, EPI_ISL_7570970, EPI_ISL_7570988, EPI_ISL_7570993 | | | |
| see above | OHSU Lab Services Molecular Microbiology | Oregon SARS-CoV-2 Genome Sequencing Center | Alec J. Hirsch; Andrew C. Adey; Benjamin N. Bimber; Brendan L. O'Connell; Brian J. O'Roak; Cierra LaBlanc; Daniel N. Streblow; Destine Krenik; Donna Hansel; Guang Fan; Rabeka Ali; Ruth V. Nichols; Sonia Acharya; William B. Messer; Xuan Qin |

| | | | |
|---|---|---|---|
| | Lab | | |
| EPI_ISL_7273927, EPI_ISL_7273967, EPI_ISL_7274134, EPI_ISL_7275370, EPI_ISL_7275609 | Ontario's COVID-19 Genomics Rapid Response Coalition | McMaster University | Ahmed Draia; Allison McGeer; Andrew G. McArthur; Angel Li; Emily Panousis; Hooman Derakhshani; Jalees Nasir; Kuganya Nirmalarajah; Michael Surette; Patryk Aftanas; Samira Mubareka; Sheridan Baker |
| EPI_ISL_7547548 | PRVKP FKUI | National Institute of Health Research and Development | Arie Ardiansyah Nugraha; Fajar Nur; Hana Apsari Pawestri; Hartanti Dian Ikawati; Kartika Dewi Puspa; Krisna Pangesti; Nelly Puspandari; Subangkit; Sulistiyohadi; Vivi Setiawaty |
| EPI_ISL_7129868, EPI_ISL_7129869 | Pandemic Response Lab - NYC | Pandemic Response Lab, R&D | Alex Carpio; Cybill del Castillo; Dylan Law; Haiping Hao; Henry Lee; Isabel Fernandez Escapa; Jon Laurent; Melissa Hopkins; Michael Hammerling; Pradeep Bugga; Shinyoung Clair Kang; Sol Rey; William Ward |
| EPI_ISL_7263932, EPI_ISL_7263933 | Pathogenic Microorganisms Variability Laboratory | Pathogenic Microorganisms Variability Laboratory | Alexander Gintsburg; Alexander Voskoboinikov; Andrei Siniavin; Andrey Pochtovyy; Artem Tkachuk; Denis Logunov; Elena Shidlovskaya; Elizaveta Divisenko; Inna Dolzhikova; Lyudmila Vasilchenko; Nadezhda Kuznetsova; Odintsova Alina; Vladimir Gushchin |
| EPI_ISL_6956014 | Pathology West - NSW Health Pathology | NSW Health Pathology - Institute of Clinical Pathology and Medical Research; Westmead Hospital; University of Sydney | Arnott A.; Draper J.; Gall M.; Martinez E.; Rockett R.; Sintchenko V.; on behalf of ICPMR |
| EPI_ISL_7173899 | Platform BIS UZA/UAntwerpen | Labo Klinische Biologie, UZA | Basil Britto Xavier; Christine Lammens; Herman Goossens; Ines Verbesselt; Jasmine Coppens; Kathleen Holemans; Marie Le Mercier; Veerle Matheeussen |
| EPI_ISL_7337575, EPI_ISL_7337577, EPI_ISL_7337600 | Port Elizabeth Laboratory | National Institute for Communicable Diseases of the National Health Laboratory Service | Amoako DG; Bhiman JN; Everatt J; Ismail A; Mahlangu B; Mnguni A; Mohale T; Ntuli N; Scheepers C |
| EPI_ISL_7135501, EPI_ISL_7135504, EPI_ISL_7263925, EPI_ISL_7263926 | Public Health Laboratory, Public Health Service Amsterdam, The Netherlands | Department of Medical Microbiology & Infection prevention, Amsterdam University Medical Centers location AMC | Akke Cornelissen; Fokla Zorgdrager; Janke Schinkel; Jelle Koopsen; Judith den Uil; Marcel Jonges; Matthijs Welkers; Menno de Jong; Robin van Houdt; Sebastien Matamoros; Sjoerd Rebers; Sylvia Bruisten; Tjalling Leenstra and Mariken van der Lubben on behalf of the Amsterdam Regional Genomic epidemiology and Outbreak Surveillance (ARGOS) consortium |
| EPI_ISL_7613143 | Public Health Ontario Laboratory | Public Health Ontario Laboratory | Aimin Li; Alireza Eshaghi; Andre Villegas; Ashleigh Sullivan; Christine Frantz; Dean Maxwell; Esha Joshi; Jared Simpson; Jennifer L Guthrie; Jonathan B Gubbay; Karthikeyan Sivaraman; Lawrence Heisler; Matthew Watson; Michael CY Li; Michael Laszloffy; Nahuel Fittipaldi; Philip Banh; Richard de Borja; Samir N Patel; Sandeep Nagra; Sandra Zittermann; Sarah Teatero; Vanessa G Allen; Yao Chen; Yogi Sundaravadanam |
| EPI_ISL_5846287, EPI_ISL_5846288, EPI_ISL_5929268 | Quest Diagnostics Incorporated | Centers for Disease Control and Prevention Division of Viral Diseases, Pathogen Discovery | A. Gerasimova; A. Perez; B. Anderson; Benjamin Rambo-Martin; Christopher Gulvick; Clinton Paden; Dakota Howard; Dhwani Batra; Duncan MacCannell; Erisa Sula; F. Lacbawan; I. Shlyakhter; Jason Caravas; K. Livingston; Kristine Lacek; L. Bernstein; M. Hua; Matthew Schmerer; P. Tanpaiboon; Peter Cook; R. Kagan; R. Owen; R. Rolando; S. Rosenthal; Scott Sammons; Shatavia Morrison; Tymeckia Kendall; Victoria Caban Figueroa; Y. Liu; Yvette Unoarumhi |
| EPI_ISL_6693526 | ROB FERREIRA LABORATORY | National Institute for Communicable Diseases of the National Health Laboratory Service | Amoako DG; Bhiman JN; Everatt J; Ismail A; Mahlangu B; Mnguni A; Mohale T; Ntuli N; Scheepers C |
| EPI_ISL_3160245 | RSUD Raden Mattaher | Eijkman Institute for Molecular Biology, National Research and Innovation Agency; Raden Mattaher Regional General Hospital | Amin Soebandrio; Edison Johar; Fery Kusnadi; Frilasita A Yudhaputri; Hidayat Trimarsanto; Iskandar Adnan; Khin Saw Myint; Lidwina Priliani; Lydia V. Panggalo; Muhammad Rezki Rasyak; Safarina G Malik; Sotianingsih; Sukma Oktavianthi; Willy Agustine; Yusnidar |
| EPI_ISL_7345201, EPI_ISL_7399058, EPI_ISL_7399078, EPI_ISL_7463969 | Respiratory Virus Unit, Microbiology Services Colindale, Public Health England | COVID-19 Genomics UK (COG-UK) Consortium | PHE Covid Sequencing Team |
| EPI_ISL_7283557, EPI_ISL_7283618 | Right to Care Zambia | Macha Research Trust | Gill CJ; Katowa B; Kwenda G; Matoba J; Mubemba B; Muleya W; Mupila Z; Mwananyanda L; Simulundu E |
| EPI_ISL_6773844, EPI_ISL_6774094, EPI_ISL_6774098, EPI_ISL_6774460, EPI_ISL_6775184, EPI_ISL_6775637, EPI_ISL_6776308, EPI_ISL_6776320, EPI_ISL_6776322, EPI_ISL_6776889, EPI_ISL_6777153, EPI_ISL_6777155, EPI_ISL_6794911, EPI_ISL_6794948, EPI_ISL_6795029, EPI_ISL_6795034, EPI_ISL_6795401, EPI_ISL_6795410, EPI_ISL_6795415, EPI_ISL_6795418, EPI_ISL_6814786, EPI_ISL_6814817, EPI_ISL_6814818, EPI_ISL_6814843, EPI_ISL_6814915, EPI_ISL_6814919, EPI_ISL_6829580, EPI_ISL_6829629, EPI_ISL_6829632, EPI_ISL_6830399, EPI_ISL_6831171, EPI_ISL_6831755, EPI_ISL_6832992, EPI_ISL_6832993 | | | |
| see above | Right to care Zambia | Macha Research Trust | Gill CJ; Katowa B; Kwenda G; Matoba J; Mubemba B; Muleya W; Mupila Z; Mwananyanda L; Simulundu E |
| EPI_ISL_7366154 | Rush University Medical Center | RIPHL at Rush University Medical Center | Alyse Kittner; Cecilia Chau; Diane Springer; Edith Perez; Felix Araujo Perez; Hannah Barbian; Joyce Houlihan; Kevin Kunstman; Laura Furtado; Marieta Hyde; Mary Hayden; Sofiya Bobrovska; Stefan Green |
| EPI_ISL_6913953, EPI_ISL_6914908, EPI_ISL_7194610 | SARS-CoV-2 testing team, National Institute of Infectious Diseases | Pathogen Genomics Center, National Institute of Infectious Diseases | Hazuka Y Furihata; Kentaro Itokawa; Makoto Kuroda; Masanori Hashino; Masumichi Saito; Naomi Nojiri; Nozomu Hanaoka; Rina Tanaka; Tsuguto Fujimoto; Tsuyoshi Sekizuka |
| EPI_ISL_1445272 | SERV DE VIG SANITARIA EPIDEMIO E CTRL DE ZOONOZES GUARUJA | Instituto Butantan / Mendelics | Antonio Jorge Martins; Bibiana Santos; Claudia Renata dos Santos Barros; David Schlesinger; Debora Botequio Moretti; Dimas Tadeu Covas; Elaine Cristina Marqueze; Elaine Vieira dos Santos; Erika Freitas; Evandra Strazza Rodrigues; Flavia Aburjaile; José Salvatore Leister Patané; João Paulo Kitajima; Luiz Carlos Junior de Alcantara; Maria Carolina Elias; Marta Giovanetti; Rafael dos Santos Bezerra; Raul Machado Neto; Ricardo Haddad; Rodrigo Tocantins Calado.; Sandra Coccuzzo Sampaio; Simone Kashima; Svetoslav Nanev Slavov; Vagner Fonseca; Vincent Louis Viala |
| EPI_ISL_6492245 | SRC VB "Vector", "Collection of microorganisms" Department. | State Research Center of Virology and Biotechnology (VECTOR) Collection of microorganisms Department | Alexander N. Shvalov; Anastasiya A. Nazarenko; Anastasiya M. Smirnova; Elena V. Gavrilova; Oleg V. Pyankov; Rinat A. Maksyutov; Sergey A. Bodnev; Tatyana V. Tregubchak |
| EPI_ISL_6512569, EPI_ISL_6512570 | SURA | Laboratorio Departamental de Salud Publica de Antioquia | Ana Victoria Valencia Duarte; Cristian Arbey Velarde Hoyos; Gloria Isabel Escobar; Idabely Betancur Ortiz; Juan P. Hernandez-Ortiz; Juan Pablo Isaza Agudelo; Maria Stella López |
| EPI_ISL_2893886 | SYNLAB GASCOGNE | CNR Virus des Infections Respiratoires - France SUD | Antonin Bal; Bruno Lina; Gregory Destras; Gwendolyne Burfin; Hadrien Regue; Laurence Josset; Martine Valette; Quentin Semanas |
| EPI_ISL_6774035 | Seberang Perai Tengah District Health Office | Institute for Medical Research, Infectious Disease Research Centre, National Institutes of Health, Ministry of Health Malaysia | Ahmad FA; Ahmad Fazilah NA; Anasir MI; Azizan MA; Kamel K; Mohamad Sukri MZ; Mohd Zawawi Z; Norhisham SN; Ramly N; Robert F; Rosli NR; Suppiah J; Thayan R |
| EPI_ISL_2439182, EPI_ISL_2439183, EPI_ISL_2439188, EPI_ISL_2801976, EPI_ISL_3411550 | Servicio Microbiología Hospital La Paz | Servicio Microbiología Hospital La Paz | Elie Dahdouh; Fernando Lázaro; Iván Bloise; Jesús Mingorance Cruz; Rubén Cáceres |
| EPI_ISL_4083348, EPI_ISL_6695068, EPI_ISL_6695069, EPI_ISL_6695071, EPI_ISL_6695072, EPI_ISL_6695092, EPI_ISL_6695109, EPI_ISL_6695110, EPI_ISL_6695221, EPI_ISL_6695235, EPI_ISL_6695248, EPI_ISL_6695258, EPI_ISL_6695275 | | | |
| see above | Servicio Virosis Respiratorias-Departamento Virología-INEI | Instituto Nacional Enfermedades Infecciosas C.G.Malbran | Avaro M.; Baumeister E.; Benedetti E.; Campos J.; Cisterna D.; Dattero ME; De Belder D.; Haim MS.; Lorenzo F.; Molina V.; Perandones C.; Poklepovich T.; Pontoriero A.; Russo M.; Sanchez Loria J.; Tuduri E. |
| EPI_ISL_2790432 | Servizo de Microbioloxía. Complexo Hospitalario de Santiago de Compostela | Servizo de Microbioloxía. Complexo Hospitalario de Santiago de Compostela | Amparo Coira; Antonio Aguilera; Carlos García_Riestra; Daniel Navarro; Gema Barbeito; Iria Roca; José Llovo; Laura Millán; Manuela Hernández; María Luisa Pérez_del_Molino; Mercedes Treviño; Rocío trastoy; Teresa Lopez_Valiño; Xosé Costa |
| EPI_ISL_7056045 | Shamir Medical Center (Asaf Harofe) | Shamir Medical Center (Asaf Harofe) | Abu Hamad Ramzia; Adina Bar Chaim; Alona Frenkel; Anna Vishnevsky; Chen Weiner; Nir Rainy; Patricia Benveniste-Lekovitz; Reut Sorek Abramovich; Yevgeni Yegorov |
| EPI_ISL_7013425, EPI_ISL_7264087, EPI_ISL_7456351, EPI_ISL_7456393, EPI_ISL_7456394, EPI_ISL_7456395, EPI_ISL_7456396, EPI_ISL_7456397, EPI_ISL_7456398, EPI_ISL_7456399, EPI_ISL_7456400 | | | |
| see above | State Laboratories Division, Hawaii State Department of Health | State Laboratories Division, Hawaii State Department of Health | Ayana Garnet; Daniel Strange; Drew Kuwazaki; Edward Desmond; Pamela O'Brien; Razvan Sultana |
| EPI_ISL_7570014, EPI_ISL_7570130 | TPMG Regional Laboratory | California Department of Public Health | Emily Smith on behalf of CDPH-COVIDNet and Invitae |
| EPI_ISL_1678096 | UT-Unified State Labs: Public Health Utah DOH | Centers for Disease Control and Prevention Division of Viral Diseases, Pathogen Discovery | Alison Laufer Halpin; Ben L. Rambo-Martin; Clinton R. Paden; Dakota Howard; Darlene Wagner; Dave Wentworth; Dhwani Batra; Jasmine Padilla; Justin Lee; Katie Dillon; Krista Queen; Kristen Knipe; Kristine Lacek; Mark Burroughs; Matthew Schmerer; Mili Sheth; Peter Cook; Sam Shepard; Sarah Nobles; Shoshona Le; Suxiang Tong; Vivien Dugan; Yvette Unoarumhi |
| EPI_ISL_7160037, EPI_ISL_7160038, EPI_ISL_7160039 | UW Virology Lab | UW Virology Lab | Alexander Greninger; Hong Xie; Keith R Jerome; Meei-Li Huang; Pavitra Roychoudhury; Pooneh Hajian; Ricardo Perez; Saraswathi Sathees; Sean Ellis; Shah Mohamed Bakhash; Tien V. Nguyen |
| EPI_ISL_2942352 | Unidad de Investigacion | Unidad de Genomica Avanzada | ; Alejandra Garcia-Gasca; Alejandra Hernandez-Teran; Alejandro Sanchez-Flores; Alfredo Herrera-Estrella; Alicia Ocaña-Mondragon; Andreu Comas-Garcia; Angel Gustavo Salas-Lais; Antonio Loza Roman; Bernardo Martinez-Miguel; Blanca Taboada; Brenda Irasema Maldonado-Meza; Bruno Gomez-Gil; Carla Ivon |

| | | | |
|---|---|---|---|
| | Medica de Yucatan (UIMY) | | Herrera-Najera; Carlos F. Arias; Celia Boukadida; Celida Duque Molina; Celida Martinez- Rodriguez; Clara Esperanza Santacruz-Tinoco; Concepcion Grajales-Muñiz; Consorcio Mexicano de Vigilancia Genomica (CoViGen-Mex). Authors (in alphabetical order): Julio Elias Alvarado-Yaah; Cristobal Chaidez-Quiroz; Daniel Fregoso-Rueda; Daniel Lira Morales; Eduardo Becerril-Vargas; Fernando Fontove-Herrera; Fidencio Mejia-Nepomuceno; Francisco Pulido; Gloria Elena Espinosa-Ayala; Gloria Maria Molina-Salinas; Gloria Vazquez; Hector Esteban Paz-Juarez; Hector Montoya-Fuentes; Helen Haydee Fernanda Ramirez-Plascencia; Irvin Gonzalez-Lopez; Jean Pierre Gonzalez; Jesus Hernandez; Joel Armando Vazquez-Perez.; Jorge Salas-Hernandez; Jose Antonio Enciso-Moreno; Jose Arturo Martinez-Orozco; Jose Esteban Muñoz-Medina; Jose de Jesus Nuñez-Contreras; Juan Bautista Chale-Dzul; Julissa Enciso-Ibarra; Luis Alberto Ochoa-Carrera; Margarita Matias-Florentino; Maria Guadalupe Santiago-Mauricio; Maria Guadalupe de Jesus Mireles-Rivera; Mario Mujica-Sanchez; Marissa Perez-Garcia; Nelly Selem-Mojica; Pavel Isa; Ricardo Ciria Merce; Ricardo Grande; Rosa Maria Gutierrez Rios; Santiago avila-Rios; Selene Zarate; Susana Lopez; Veronica Mata-Haro; Victor Eduardo Garcia-Arias; Victor Hugo Borja-Aburto |
| EPI_ISL_5887477 | Universidad San Sebastian | "Facultad de Ciencias de la Vida, UNAB" | "Claudio Meneses; Ariel Orellana"; Claudio Olmos; Daniel Leon; Dayan Sanhueza; Eduardo Castro; Gonzalo Campaña; Macarena Bastias; Paola Pidal; Ricardo Yusta; Sebastian Wolter; Susana Saez; Víctor Monreal; Waldo Diaz |
| EPI_ISL_5858827, EPI_ISL_5858842 | Universidad del Valle, LDAB-Laboratorio de Diagnostico de Agentes Biologicos | Universidad del Valle | Andres Castillo; Beatriz Parra & Programa Nacional de Caracterización Genómica de SARS-CoV-2; Diana López-Alvarez; Erica M. Aristizabal; Flor Saa; Melissa Solarte; Nelson Rivera Franco |
| EPI_ISL_2091066 | University Hospitals Translational Laboratory (UHTL), University Hospitals | University Hospitals Translational Laboratory (UHTL), University Hospitals | Alouani, D.; Sadri, N.; Song, X. |
| EPI_ISL_2213596 | University Hospitals of Geneva, Laboratory of Virology | HUG, Laboratory of Virology and the Health2030 Genome Center | Ana Rita Goncalves; Deborah Penet; Emmanouil Dermitzakis; Henri Pegeot; Ioannis Xenarios; Keith Harshman; Laurent Kaiser; Lorenzo Cerutti; Melyssa Elies; Samuel Cordey |
| EPI_ISL_6510847, EPI_ISL_6510855 | Università degli Studi di Perugia, Perugia, Italy | Istituto Superiore di Sanità (ISS) | Alessandra Lo Presti; Angela Di Martino; Antonella Mencacci; Barbara Camilloni; Luigina Ambrosio; Manuela Marra; Marco Crescenzi; Maria Carollo; Paola Stefanelli |
| EPI_ISL_7136300, EPI_ISL_7136771, EPI_ISL_7336152 | Utah Public Health Laboratory | Utah Public Health Laboratory | Erin L. Young; John Arnn; Kelly F. Oakeson; Olinto Linares-Perdomo; Pooja Gupta; Tom Iverson |
| EPI_ISL_7398758 | Vichaivej International Hospital Nongkhaem | National Institute of Health, Department of Medical Sciences, Ministry of Public Health, Thailand | Archawin Rojanawiwat; Ballang Uppapong; Beth Skaggs; Donlaya Maunplueng; Kazuhisa Okada; Natchaya Khiadsang; Nuttida Thongpramul; Pakorn Piromtong; Pilailuk Akkapaiboon Okada; Piroon Jenjaroenpun; Pongpun Sawatwong; Prapat Suriyaphol; Sirikanda Wimol; Siripaporn Phuygun; Sittiporn Parnmen; Supakit Sirilak; Suratchana Mitrat; Thanutsapa Thanadachakul; Thidathip Wongsurawat |
| EPI_ISL_7372207 | Viollier AG | Department of Biosystems Science and Engineering, ETH Zürich | Andrea Patrignani; Andreia Cabral de Gouvea; Catharine Aquino; Chaoran Chen; Christiane Beckmann; Christoph Noppen; Daniel Ehrsam; Doris Popovic; Griffin White; Isabel Stürmer; Ivan Topolsky; Jay Tracy; Kim Philipp Jablonski; Lara Fuhrmann; Laura Neff; Lennart Opitz; Louis du Plessis; Maria Domenica Moccia; Maurice Redondo; Niko Beerenwinkel; Olivier Kobel; Ralph Schlapbach; Sarah Nadeau; Simon Grüter; Tanja Stadler; Timothy Sykes |
| EPI_ISL_6795847, EPI_ISL_6795848, EPI_ISL_6825393, EPI_ISL_6825395 | ZARV/NHLS, Department Medical Virology, University of Pretoria | CERI, Centre for Epidemic Response and Innvoation, Stellenbosch University and KRISP, KZN Research Innovation and Sequencing Platform, UKZN. | Adriano Mendes; Amoaka D; Amy Strydom; Arisha Maharaj; Bester P; Bhiman J; Engelbrecht S; Everatt J; Goedhals D; Hardie D; Hsiao M; Iranzadeh A; Lessells R; Makatini Z; Maponga T; Mdlalose N; Micheala Davids; Mlisana K; Moir M; NGS-SA (Scheepers C; Naidoo Y; Nyaga M) Giandhari J; Oluwakemi M; Pillay S; Preiser W; Ramphal U; Ramphal Y; San JE; Sim Mayaphi and Marietjie Venter; Tegally H; Tshiabuila D; Venter M; Wilkinson E; Williamson C; de Oliveira T; von Gottberg A |
| EPI_ISL_6136195 | unknown | Noblis | Katharine Jennings; Lauren Brinkac; Lee F. Kolakowski; Minh Tran; Shane Mitchell; Shiela Diepold; William M. Nelson |
